# Exploiting pleiotropy to enhance variant discovery with functional false discovery rates

**DOI:** 10.1101/2024.09.24.24314276

**Authors:** Andrew J. Bass, Chris Wallace

## Abstract

The cost of acquiring participants for genome-wide association studies (GWAS) can limit sample sizes and inhibit discovery of genetic variants. We introduce the surrogate functional false discovery rate (sfFDR) framework which integrates summary statistics of related traits to increase power. The sfFDR framework provides estimates of FDR quantities such as the functional local FDR and *q*-value, and uses these estimates to derive a functional *p*-value for type I error rate control and a functional local Bayes’ factor for post-GWAS analyses (e.g., fine mapping and colocalization). Compared to a standard analysis, sfFDR substantially increased power (equivalent to a 60% increase in sample size) in a study of obesity-related traits from the UK Biobank, and discovered eight additional lead SNPs near genes linked to immune-related responses in a rare disease GWAS of eosinophilic granulomatosis with polyangiitis. Collectively, these results highlight the utility of exploiting related traits in both small and large studies.

## 1 Introduction

Genome-wide association studies (GWAS) provide a wealth of genetic data to understand the aetiology of human diseases. In a GWAS, the discovery of genetic variants requires an adequate sample size to represent the population and maximize statistical power. While increasing sample size increases variant discovery, the sample size is often limited by the cost or availability of participants, particularly in the case of low frequency or rare diseases.

Given such sample size constraints, an alternative approach is to leverage the ubiquitous genetic correlations (i.e., pleiotropy) between related traits to improve variant discovery [1–4]. One strategy is to use GWAS summary statistics of related traits within a conditional false discovery rate (cFDR) framework to increase power [5]. While a typical GWAS analysis aims to control the probability of at least one false discovery (defined as a variant that does not tag a causal variant), the cFDR approach is more liberal in that it controls the expected proportion of false discoveries (i.e., the FDR [6]). Previous work on the cFDR has shown a substantial increase in power when incorporating GWAS summary statistics of related traits compared to a standard GWAS [5, 7, 8], and thus has been applied in GWAS to enhance discovery of variants (see, e.g., [9–12]). However, the utility of cFDR approaches are limited due to computational cost and strict assumptions of independence between related traits. Although there are other general FDR procedures that can integrate informative data [13–16], it is unclear how to appropriately incorporate GWAS summary statistics while accommodating for dependence due to linkage disequilibrium (LD). Therefore, current approaches can not fully leverage pleiotropy from multiple related traits to increase power. More generally, the familiar guarantees of familywise error rate (FWER) control has been a barrier to widespread adoption of FDR methods in GWAS, even though the FDR can substantially increase the number of discoveries in genomics [17].

To address these challenges, we develop a novel method that integrates multiple sets of GWAS summary statistics within the functional FDR (fFDR) framework [15]. The fFDR framework was primarily designed for genomic studies and incorporates a single informative variable (e.g., epigenetic or per-gene read depth) when constructing FDR quantities of interest, such as the functional *q*-value (a measure of significance in terms of the positive FDR [17, 18]) and local FDR (a posterior error probability [19, 20]). Our proposed method, surrogate functional FDR (sfFDR), adapts the fFDR to leverage informative data from multiple sets of GWAS sum-mary statistics while accommodating for LD. Importantly, sfFDR is a computationally efficient approach and does not assume independence between the related GWAS traits. We also derive a new quantity, the functional *p*-value, that incorporates the GWAS summary statistics and can be interpreted like a standard *p*-value familiar to GWAS practitioners. Finally, we show how functional local Bayes’ factors can be calculated from the functional local FDR, allowing a range of post-GWAS analyses to incorporate GWAS summary statistics such as functional fine mapping and colocalization.

We apply sfFDR to both small and large sample size GWAS studies to illustrate the power improvements compared to a standard GWAS analysis. We first perform comprehensive simulations to evaluate and compare sfFDR to three general FDR methods extended to our setting. We then demonstrate the power improvements in a study of obesity-related traits from the UK Biobank. Finally, we apply sfFDR to a rare disease GWAS of eosinophilic granulomatosis with polyangiitis (EGPA) and use GWAS summary statistics from related traits (asthma and eosinophil count) to substantially increase power compared to a standard GWAS analysis. We also show how estimates of the functional local FDR can be used to perform functional fine mapping in the EGPA study and thus help identify the causal locus within a genetic region (assuming a single causal locus).

## 2 Results

### 2.1 Overview

We briefly review the motivation behind the sfFDR framework (see Methods for additional details). Consider a GWAS for some trait of interest, referred to as the “primary” GWAS, where a *p*-value is calculated on a SNP-by-SNP basis to assess statistical significance. In a typical analysis, the set of SNPs below a genome-wide significance threshold (e.g., *p <* 5 *×* 10^*−*8^) are classified as statistically significant where each SNP is treated equally likely *a priori* to be truly null. However, there is often an abundance of SNP-level information available that can alter our prior belief about whether a SNP is more or less likely to be associated with the trait of interest. In particular, a valuable source of SNP-level information is from publicly available GWAS summary statistics, where traits with similar genetic architecture can be integrated into the significance analysis to improve power.

Our approach, sfFDR, leverages one or several sets of informative GWAS summary statistics within an extended version of the functional FDR framework [15] to improve the power of the primary GWAS (Figure 1). Given *p*-values from the primary GWAS and one or more informative GWAS, ***z***, we first identify a LD-independent subset of SNPs. Using the LD-independent SNPs, we estimate the functional local FDR which requires modeling the functional proportion of truly null hypotheses, *π*_0_(***z***), and the conditional density, *f* (*p* | ***z***). We estimate *π*_0_(***z***) using a generalized additive model (GAM) and *f* (*p* | ***z***) nonparametrically where we use a surrogate variable approximation—the ranked estimated *π*_0_(***z***) values—that circumvents difficulties with higher dimensional density estimation. The functional local FDR of the left-out dependent SNPs are then predicted from the model fit of *π*_0_(***z***) and *f* (*p* | ***z***). With the estimated functional local FDRs, the functional *q*-values (referred to throughout as *q*_*f*_ -value) are constructed for each SNP and measure significance in terms of the positive FDR (pFDR; closely related to FDR [20]). Intuitively, the *q*_*f*_ -value is the minimum probability that a SNP is null given that it is classified as statistically significant (i.e., the “Bayesian posterior type I error” [20]).

**Figure 1:**
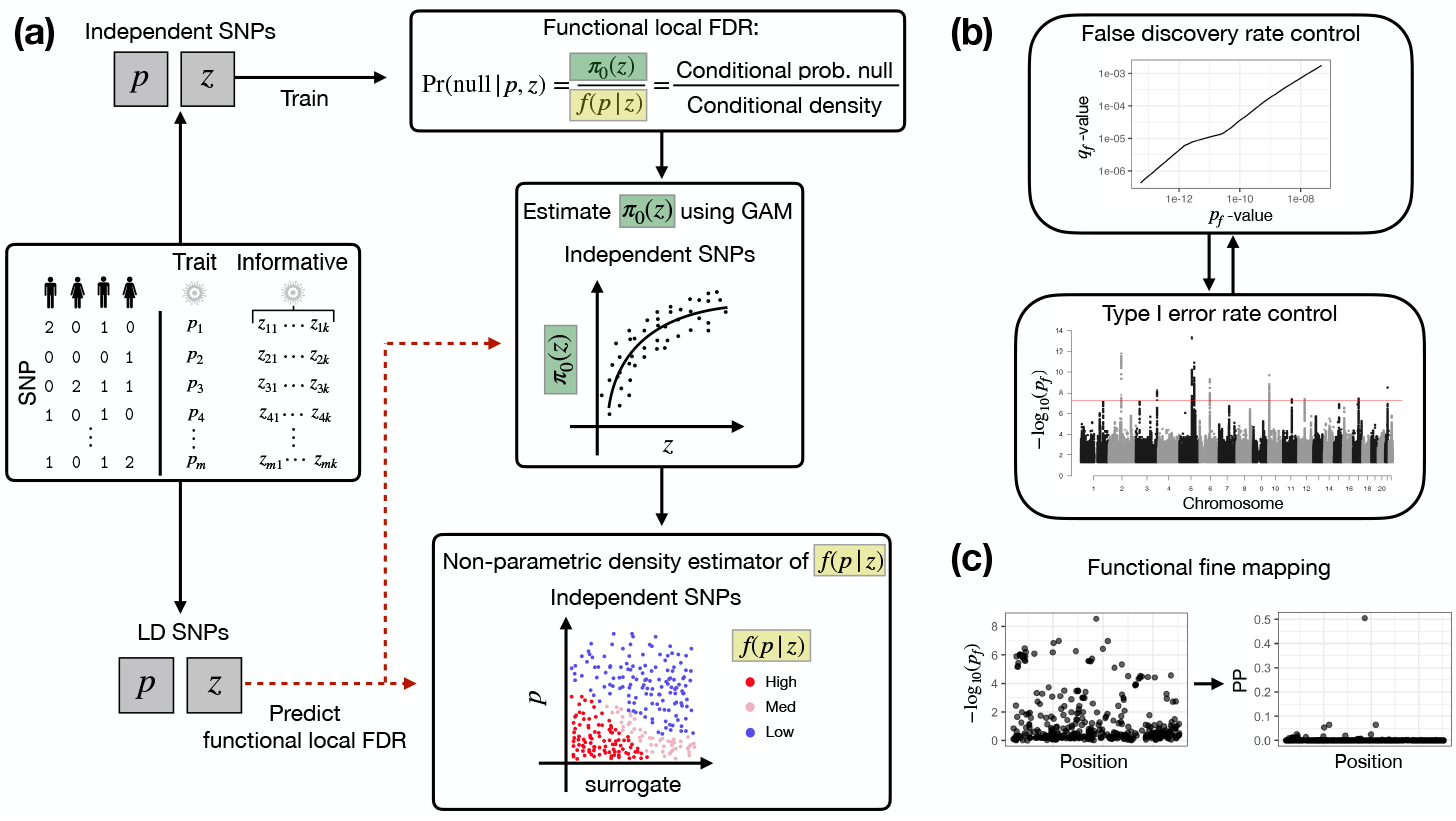
Overview of the surrogate functional false discovery rate (sfFDR) framework. (**a**) Estimate the functional local FDR of the primary GWAS *p*-values given a set of informative summary statistics. The functional local FDR values are used for (**b**) estimating the functional *q*-value (*q*_*f*_ - value) and functional *p*-value (*p*_*f*_ -value) to control the FDR and family-wise error rate, respectively, and (**c**) functional fine mapping.

The FDR quantities estimated by the sfFDR framework support a range of analyses for GWAS data. In particular, we use the FDR quantities to derive a functional *p*-value (referred to throughout as *p*_*f*_ -value), allowing practitioners to control the FWER while incorporating SNP-level information. We also use the functional local FDR to derive functional local Bayes’ factors, enabling post-GWAS analyses such as functional fine mapping to help identify the causal variant in a region (assuming a single causal variant).

### 2.2 Evaluating the sfFDR framework

We performed comprehensive simulations to evaluate the sfFDR framework in two settings (Methods). The first setting simulates independent SNPs to allow comparison with other FDR approaches while the second generates regions of LD to simulate GWAS data. Since one of our applications is a rare disease study, we focus on simulating data to reflect the challenging scenario expected in studies of low sample sizes, i.e., the genetic signal is sparse.

We simulated the *p*-values for 150,000 independent SNPs in a primary study and three informative studies. The signal strength of the studies (i.e., statistical power) was varied as “High,” “Medium,” and “Low.” The informative studies overlapped (shared non-null SNPs with the primary study) with randomly chosen values between 1.25% and 2.50% of the total number of SNPs. At the overlapping tests, the informative studies impacted both the prior probability of a SNP being null and the alternative density of the *p*-values with an effect size strength of “Large,” “Moderate,” and “None.”

We find that the estimated *q*_*f*_ -values control the FDR at level 0.01 in all settings (Figure S1), even when the informative traits provided no information on the primary trait. Furthermore, the estimated *q*_*f*_ -values have similar power to the oracle values (i.e., the true *q*_*f*_ -values) and substantially improved power compared to the standard *q*-values [18] which were calculated from the qvalue package [21] and do not use the informative studies (Figure 2a,S2). In general, as the primary or informative studies power increases, or the effect size strength of the informative studies is larger, the more information sfFDR uses to increase power. For example, when the power of the informative studies is “High,” the power of the primary study is “Medium,” and the effect size strength is “Large,” the average number of discoveries from the *q*_*f*_ -value is 241 at a target FDR of 0.01 which is much larger than the standard *q*-value (94.5). In the same example, when the power of the informative studies is “Low,” the number of discoveries decreases (131) as expected but is still larger than the standard *q*-value (67.6).

**Figure 2:**
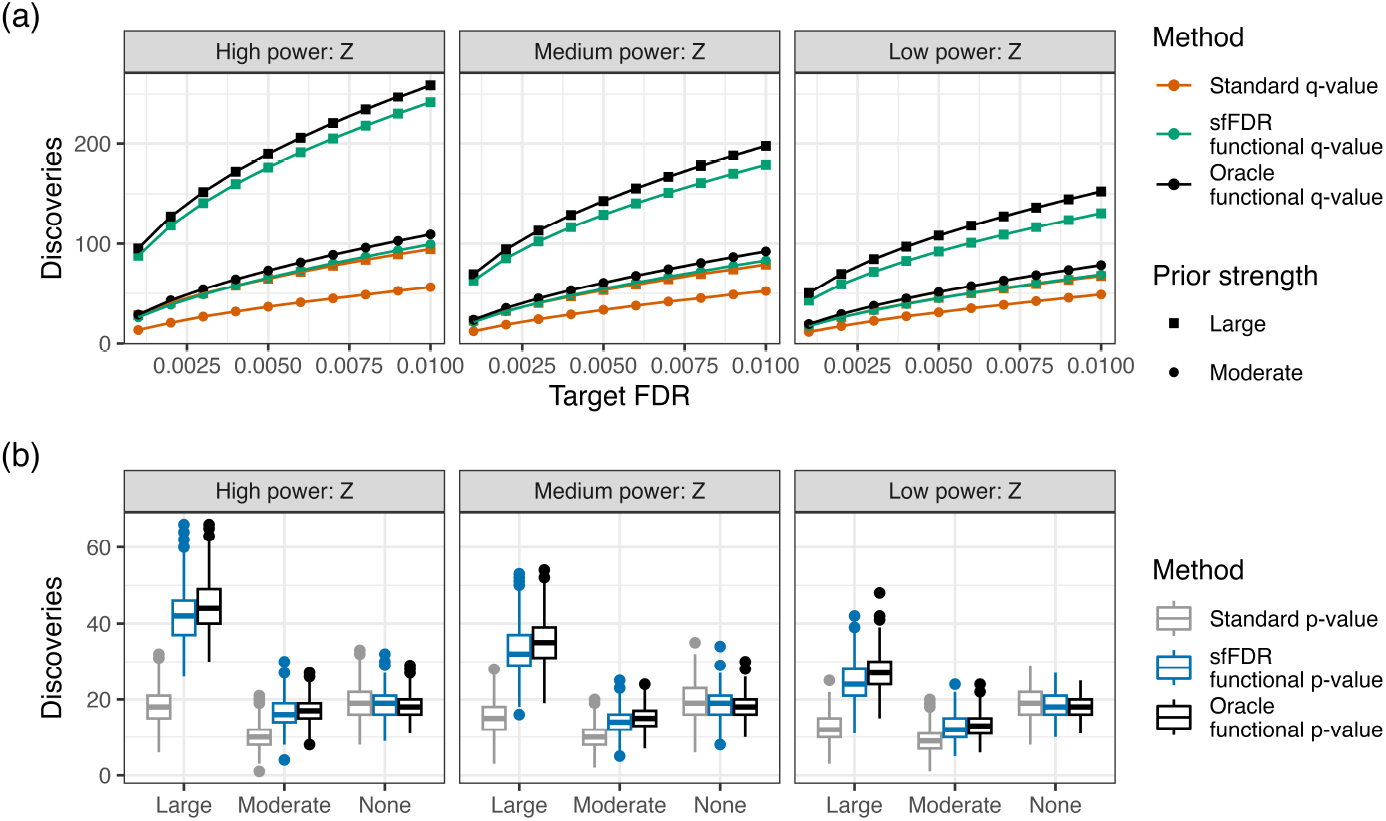
Simulation results for the sfFDR framework in the independent SNP simulation study when the primary study power is “Medium.” **(a)** The average number of discoveries as a function of the target false discovery rate (FDR) using the standard *q*-value (dark orange), functional *q*-value from sfFDR (green), and oracle functional *q*-value (black). **(b)** The number of discoveries using the standard *p*-value (grey), functional *p*-value from sfFDR (blue), and oracle functional *p*-value (black) at a genome-wide significance threshold of 5*×*10^*−*8^. We varied the power of the informative studies (columns) and the effect size strength of the informative studies (top plot: shape; bottom plot: x-axis). There were a total of 500 replicates at each setting.

We compared the sfFDR framework to other FDR procedures that can incorporate multiple informative variables, namely, AdaPT [14], CAMT [16], and an estimator by Boca et al. (2018; referred to as the “Boca-Leek” method) [13]. Overall, we find that these methods provide control of the FDR (Figure S1), although the FDR is inflated for CAMT when the primary study power is “Low” (Figure S3). Furthermore, sfFDR and CAMT have comparable power and outperform AdaPT and Boca-Leek across a range of small FDR thresholds (Figure S3). We then compared estimates of the proportion of truly null hypotheses and find that CAMT is anti-conservative (predicts more non-null SNPs than exist), AdaPT and Boca-Leek are slightly anti-conservative, and sfFDR is conservative (Figure S4). Note that a conservative estimator is preferred compared to an anti-conservative one because it does not overestimate the amount of signal which can lead to an inflated FDR.

The estimated *q*_*f*_ -value and proportion of truly null tests are then used to construct the *p*_*f*_ - value in the sfFDR framework. We find that the estimated *p*_*f*_ -value controls the type I error rate at a significance threshold of 1*×*10^*−*4^ in the independent SNP simulations (Figure S5). We also evaluated the number of discoveries at a genome-wide significance threshold of 5 *×* 10^*−*8^ and compared it to the standard *p*-values (i.e., the original *p*-values) and the oracle *p*_*f*_ -values (i.e., the true *p*_*f*_ -values; Figure 2b,S6). We find that the number of discoveries from the estimated *p*_*f*_ -values is close to the oracle *p*_*f*_ -values in all settings. As expected, the power improvements from the *p*_*f*_ -value compared to the standard *p*-value depend on the primary and informative studies power along with effect size strength. For example, the higher the power of the primary and/or informative studies coupled with a larger effect size strength, the larger the increase in the number of detections from the *p*_*f*_ -value.

Finally, we assessed control of the type I error rate and FDR in the dependent SNP setting. We first randomly assigned each independent SNP an LD block size based on the empirical distribution from the UK Biobank (Methods). Given the block size, we then duplicated the *p*-values for the primary and informative studies so that the LD block was perfectly correlated. While this represents an unrealistic scenario, it is a deliberately challenging setting to evaluate estimates in the sfFDR framework. Even under such an extreme case, we find that the estimated *p*_*f*_ -value and *q*_*f*_ -value from sfFDR controls the type I error rate (Figure S7) and FDR (Figure S8, S9), respectively. As expected, due to LD, the observed type I error rate and FDR variability is larger compared to the independent SNPs case. Nevertheless, the estimated *p*_*f*_ -value has a similar variability to the standard *p*-value.

### 2.3 sfFDR increases power in GWAS of BMI from UK Biobank

In order to investigate the behavior of sfFDR in real data, we split 390,600 unrelated individuals from the UK Biobank into two separate data sets of equal size (Section 4.6): the first (the primary study) was used to detect genetic associations for body mass index (BMI) while the second (the informative study) was used to provide *p*-values for body fat percentage (BFP), triglycerides, and cholesterol as informative traits. We then conducted a sfFDR analysis of BMI informed by the three obesity-related traits and compared it to a standard GWAS analysis of BMI.

We downsampled the primary study to examine the behavior of sfFDR at different sample sizes. We find that the number of discoveries from sfFDR is substantially larger than the standard GWAS analysis across a range of sample sizes (Figure 3a). Furthermore, we find that the discoveries made with the *p*_*f*_ -values from sfFDR are nearly all a subset of those made by meta-analysis of both data sets (BMI only; Figure S10), suggesting that the additional discoveries are a subset of those that would be found by increasing the sample size. Thus, these results demonstrate the potential of sfFDR to substantially increase the power in GWAS studies by leveraging related traits.

**Figure 3:**
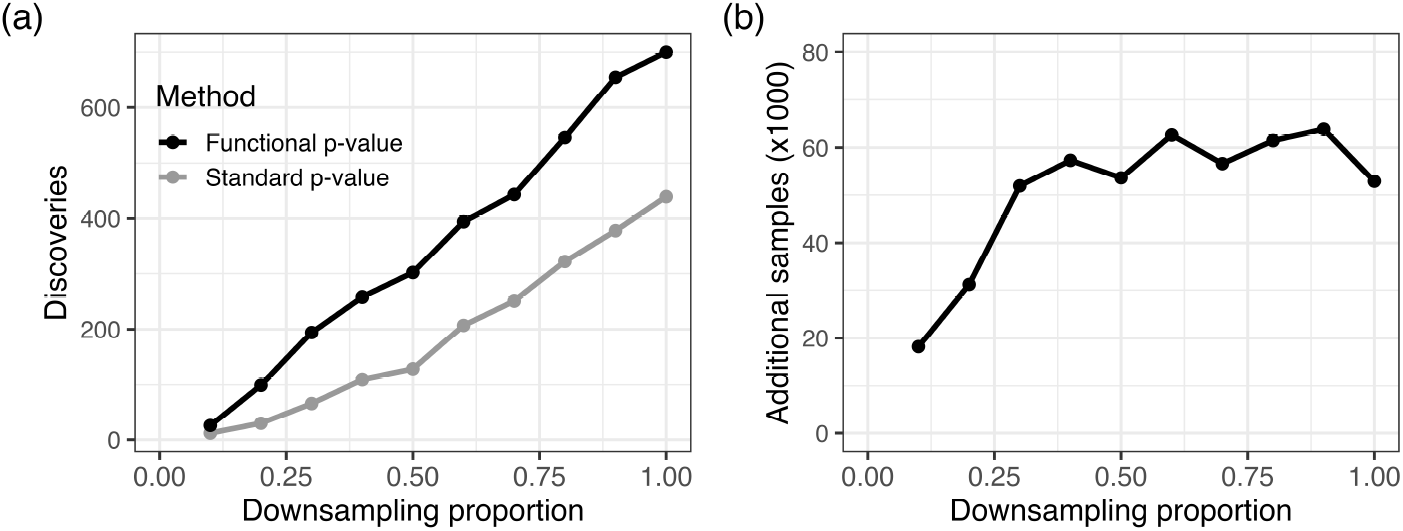
Comparing the functional *p*-value from sfFDR to the standard *p*-value from a GWAS analysis of BMI in the UK Biobank. (a) The number of discoveries as a function of the proportion of the study sample size (i.e., downsampling proportion) at a significance threshold of 5 *×* 10^*−*8^ and (b) the additional samples required for the GWAS to detect the same number of discoveries as sfFDR. We split the UK Biobank data into primary and informative studies, each with a sample size of 190,300. The standard *p*-values are calculated from the primary study (BMI) while the functional *p*-values also leverage summary statistics of additional obesity-related traits (BFP, cholesterol, and triglycerides) from the informative study.

The improvements in statistical power from sfFDR can also be translated in terms of sample size (Figure 3b). At each downsampling proportion, we predicted the sample size needed for the standard *p*-values to detect the same number of discoveries as the *p*_*f*_ -values from sfFDR. The difference in sample sizes between these values is the number of additional samples required for the standard *p*-value to match the discoveries found by the *p*_*f*_ -value. We find that the number of additional samples required is quite substantial at each downsampling proportion. For example, at a downsampling proportion of 0.4 (sample size of 76,120), the number of additional samples required is approximately 57,000 (a *∼*75% increase in sample size). Averaged across all downsampling proportions, we find that the power improvements from sfFDR equate to a *∼*60% increase in sample size.

To assess sfFDR under a scenario where the conditioning traits are uninformative, we permuted trait values in the informative study 10 times to generate traits that were uncorrelated with the primary study trait (BMI) while conserving the between trait correlations. We find that the *p*_*f*_ -value from sfFDR does not find more discoveries compared to the standard *p*-value (Figure S11) and tends to underestimate the true value at small *p*-values (i.e., conservative). We note that a conservative estimator is desired in the null setting compared to an anti-conservative one which would inflate the type I error rate. Furthermore, this behavior is expected due to the conservative estimate of the functional proportion of truly null hypotheses from sfFDR (see Methods). In general, since the *p*_*f*_ -value is incorporating additional non-informative data, it is a less accurate (or “noisy”) estimator of the standard *p*-value. Importantly, we find that using uninformative traits does not systematically inflate the significance of the *p*_*f*_ -values in real data, agreeing with our simulation results.

### 2.4 sfFDR reveals new genetic variants in the EGPA study

The sfFDR framework offers potential benefits in the rare disease setting because it is difficult and costly to acquire additional samples to improve power. As such, we applied the sfFDR framework to a GWAS of EGPA (676 cases and 6,809 controls) [12], which is a rare inflammatory disease with a prevalence of around 45.6 per 1,000,000 individuals in the UK [22]. The aetiology of EGPA unknown but is often characterized with other clinical features such as asthma and low eosinophil count [12]. Therefore, these traits are strong candidates to increase power in the EGPA study. We used a publicly available GWAS of childhood-onset asthma (13,962 cases and 300,671 controls) [23], adult-onset asthma (26,582 cases and 300,671 controls) [23], and eosinophil count (172,275 individuals) [24] as our informative studies. After removing non-overlapping SNPs between EGPA and the informative traits, there were a total of 8,195,277 SNPs used within the sfFDR framework (Section 4.7).

We first evaluated the behavior of the sfFDR framework on EGPA with a set of unrelated traits. Using the permuted null obesity-related traits (unassociated with EGPA) from the UK Biobank analysis, we find that the estimated *p*_*f*_ -value from sfFDR tends to be slightly larger than the standard *p*-value (Figure S12). Thus, similar to the above the BMI study, the *p*_*f*_ -value from sfFDR conservatively estimates the standard *p*-value for non-informative traits. Since the permuted traits do not have any association signal, we also used the original traits (i.e., unpermuted) as a set of non-null unrelated traits. On this single realization, the estimated *p*_*f*_ -value may be smaller than the standard *p*-value, but on average tends to be slightly larger (Figure S13). Importantly, the estimated *p*_*f*_ -values do not find any newly significant SNPs at the genome-wide significance threshold. Thus, non-informative data does not inflate the type I error rate in the rare disease setting.

We then applied the sfFDR framework to the EGPA study using the EGPA-informative traits (computational time was *∼*5.40 minutes on a single core of a Apple M3 processor) and find a substantial increase in the number of discoveries compared to the standard *p*-values (Figure 4, 5). We first note that the prior probability of a SNP being null for EGPA varies as a function of the informative traits *p*-values, suggesting a shared genetic architecture between traits (Figure 4a). Furthermore, as a function of significance threshold, the *p*_*f*_ -values from sfFDR find substantially more discoveries than the standard *p*-values (Figure 4b-c). For example, at the genome-wide significance threshold, there 226 discoveries using the *p*_*f*_ -values and 15 discoveries using the standard *p*-values. Of those discoveries, sfFDR identified ten lead SNPs (i.e., independent associations) instead of two by a standard GWAS analysis (Table 1). One feature of the sfFDR framework is that the *p*_*f*_ -value can be mapped to the *q*_*f*_ -value to control the FDR (Figure 4d). At the genome-wide significance threshold, we find that the estimated *q*_*f*_ -value is 1.75 *×* 10^*−*3^, which implies that there are 0.39 expected false discoveries (defined as a significant SNP that does not tag a causal SNP) in our discovery set of 226 SNPs. Thus, the mapping to a FDR analysis allows the practitioner to choose a data-adaptive significance threshold to control the expected number of false discoveries that they are willing to incur in their analysis.

**Table 1:**
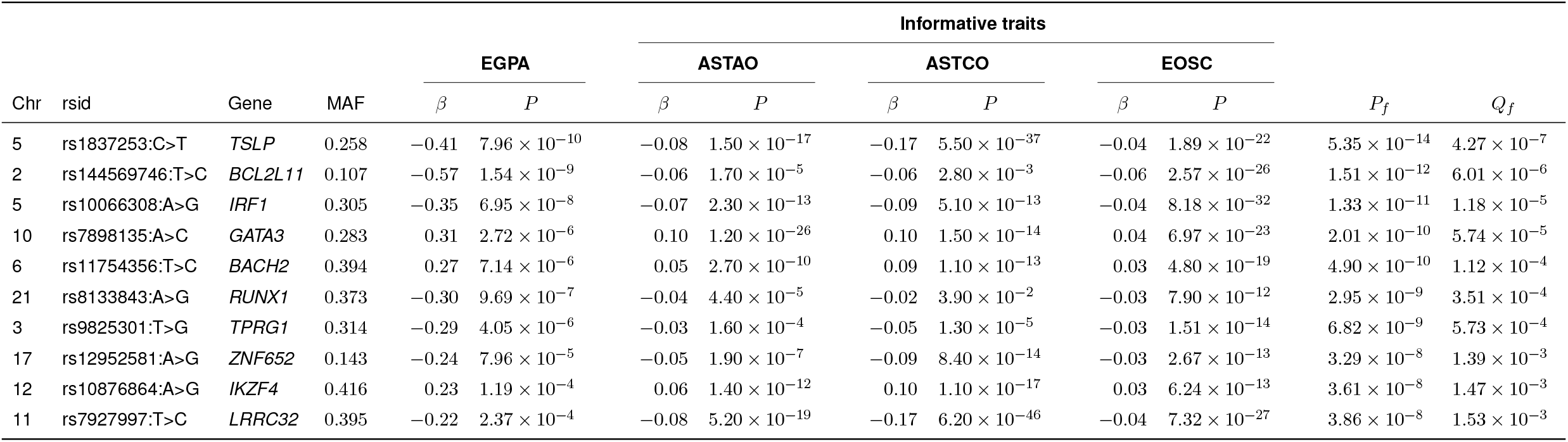
Functional *p*-values (*P*_*f*_) and *q*-values (*Q*_*f*_) of the lead SNPs from the EGPA analysis. The SNP identifiers are given as rsid:reference_allele>effect_allele. The informative traits were adult-onset asthma (ASTAO), childhood-onset asthma (ASTCO), and eosinophil count (EOSC).

**Figure 4:**
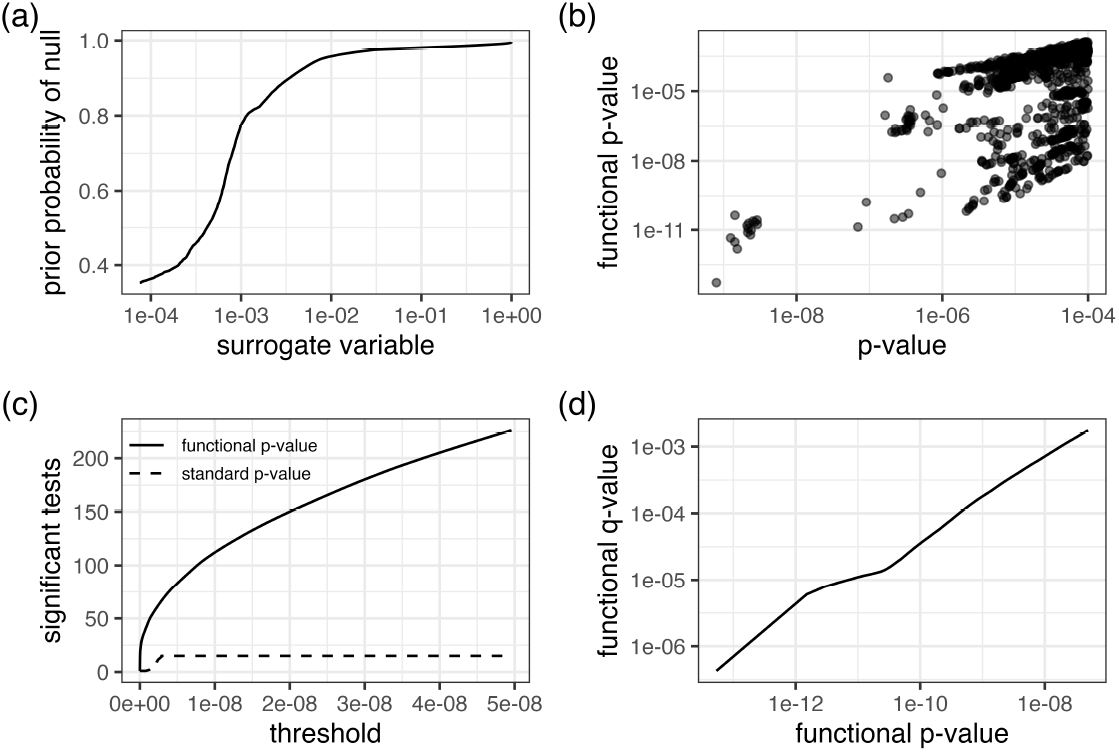
Significance results for the EGPA study. (a) The prior probability of a test being null as a function of the surrogate variable; (b) The functional *p*-value from sfFDR versus the standard *p*-value of the study; (c) The number of significant tests at various *p*-value thresholds for the functional and standard *p*-values. (d) The functional *q*-value versus functional *p*-value relationship. The above plot shows SNPs with standard *p*-values below 1 *×* 10^*−*4^ in (a)-(b) and functional *p*-values below 5 *×* 10^*−*8^ in (c)-(d).

**Figure 5:**
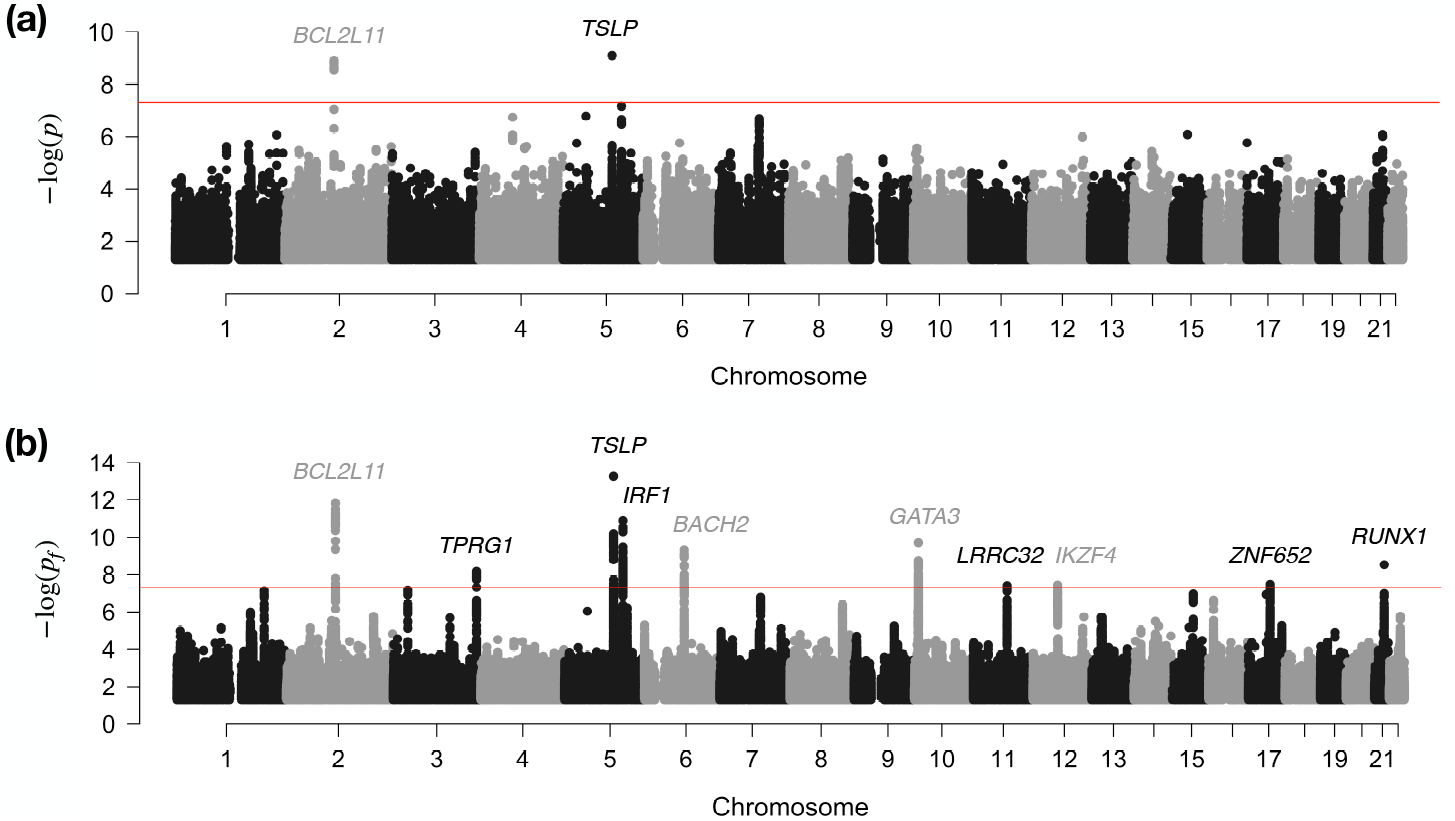
Manhattan plot of the **(a)** standard *p*-values and the **(b)** functional *p*-values (*p*_*f*_ -values) from sfFDR in the EGPA study. The red line represents the genome-wide significance threshold of 5 *×* 10^*−*8^. The lead SNPs were assigned to the nearest genes. Note that *p*-values below 0.05 are removed from the plot.

We focus our analysis on ten lead SNPs with a *p*_*f*_ -value below the genome-wide significance threshold (Table 1). After assigning SNPs to the nearest gene, we find that the original analysis with the standard *p*-values only identified two lead SNPs near *BCL2L1* and *TSLP* while the *p*_*f*_ -values from sfFDR identified eight additional genes. At these genes, the lead SNPs are either intergenic (*GATA3*), intronic (*BACH2, BCL2L11, IRF1, RUNX1, TPRG1*, and *ZNF652*), or upstream (*IKZF4, LRRC32*, and *TSLP*). Furthermore, the direction of the effect size is consistent across EGPA and the informative traits at these lead SNPs, even though the direction of the effect size is not used by the sfFDR framework.

Many of the new discoveries found by sfFDR are implicated in immune-related processes. For example, *ABI3* (161 kb from rs12952581) and *GATA3* have been linked to eosinophil counts and asthma [25], respectively, as well as *LRRC32* which encodes the eosinophilic esophagitis-associated TGF-*β* membrane binding protein *GARP*. Additionally, *IRF1* encodes a protein that activates genes involved in pro-inflammatory regulation and has been associated with childhood allergic asthma [26] where it may also have sex-specific effects [27]. Interestingly, *RUNX1* may be a prognostic marker for some cancers [28,29], and there is evidence that the *RUNX1* transcription factor is involved with Th2 cell differentiation (key for the activation of eosinophils) by decreasing *GATA3* expression [30].

One standard post-GWAS analysis is fine mapping. We fine mapped each associated region using a standard single causal variant approach with either the functional local Bayes’ factor estimated by sfFDR or the approximate Bayes’ factor (Section 4.4) [31]. Of the two genome-wide significant regions identified by the standard *p*-values (i.e., *TSLP* and *BCL2L11*), we find 1 and 14 SNPs in the 95% credible set without incorporating informative data compared to 1 and 13 SNPs using sfFDR, respectively. When extended to all the regions found by sfFDR, we find that credible sets are smaller in 7 cases (substantially in 5 cases), unchanged in 1 and larger in 2, and so a smaller credible set size is not guaranteed (Table S1). We also calculated the proportion of SNPs in the sfFDR credible sets that overlap with the credible sets of the informative traits (Table S2). Overall, we find that the sfFDR credible sets strongly overlap with the informative traits (most with eosinophil count) except at the locus in RUNX1 where only 7.70% and 8.10% of the SNPs overlap with the credible set for adult-onset asthma and childhood-onset asthma, respectively.

## 3 Discussion

We proposed a new approach, surrogate functional FDR (sfFDR), to improve power in a GWAS by leveraging summary statistics of related traits. sfFDR extends the fFDR framework [15] to integrate multiple sets of GWAS summary statistics while accommodating for LD. Although we find that sfFDR can exploit pleiotropy to substantially enhance discovery of genetic variants, FDR approaches have not been widely adopted by the GWAS community despite being commonly used in eQTL mapping. Instead, perhaps due to the abundance of non-reproducible results in earlier candidate gene studies, the preference is to control the FWER in a standard GWAS analysis. Therefore, to help GWAS practitioners leverage the power improvements from functional FDR quantities, we derived the functional *p*-value which has a standard *p*-value interpretation and can be used in a FWER-controlling procedure while incorporating informative data.

The sfFDR framework allows for a range of significance analyses in a GWAS. More specifically, sfFDR provides estimates of the functional *q*-value (a significance measure in terms of the pFDR) and the functional *p*-value (a significance measure in terms of the type I error rate). These quantities can be used to map between an FDR threshold and FWER threshold to provide an interpretation for the set of SNPs deemed statistically significant. This is useful for interpreting genetic findings in a GWAS and, more generally, as a data-adaptive way to explore the impact of false discoveries instead of an automatic application of a fixed genome-wide significance threshold. Another FDR quantity estimated by sfFDR, the functional local FDR, provides a simple way to calculate functional local Bayes’ factors which are key quantities in many post-GWAS analyses. We used it here to perform functional fine mapping under a single causal variant assumption, but it could also be used to enhance colocalization analysis using the coloc approach [32, 33].

Our simulation results have implications for the design of pleiotropy-informed significance analyses. As expected, the power improvements with sfFDR increased whenever the study power increased, for both the primary and informative study. As such, practitioners should identify informative traits that are high-powered from large GWAS studies. Fortunately, there is a large collection of GWAS summary statistics in publicly available repositories for thousands of complex traits (see, e.g., [3, 34]), although selecting the informative traits *a priori* will require careful consideration to avoid model selection (and fitting) problems. While our method can incorporate many informative studies, handling a very large number may require dimensionality reduction (e.g., principal component analysis [35] or sliced inverse regression [36]), variable selection, or regularization for stable model fitting in sfFDR. Finally, it is worth noting that our approach is not immune to sources that may bias summary statistics such as ancestry [37] or non-random sampling with respect to the reference population (e.g., participation bias [38]). In particular, if ancestry is unaccounted for in both the primary and informative studies, then there is a risk that ancestry-informative SNPs could be elevated by sfFDR. Therefore, it is important to only consider studies that adopt robust analytical strategies.

There are a few important observations when applying the sfFDR framework to GWAS data. First, estimating the prior probability of the null hypothesis (or functional proportion of truly null hypotheses) using a GAM requires specifying the relationship between the probability of a SNP being null and the informative traits. In this work, we used a natural cubic spline to flexibly model this relationship but knots have to be carefully chosen at locations where SNPs from the alternative hypothesis are likely to be located (i.e., small *p*-values). Our software includes a user-friendly function to help practitioners construct such design matrices when fitting a GAM or general linear model. Second, while we found the surrogate variable based on the functional proportion of truly null hypotheses performed well in this study, it is possible that there may be better surrogate variable choices or the nonparametric density estimation could be extended to incorporate multiple variables [15]. Third, since it is not possible to distinguish whether a (tagged) SNP is a true discovery or is capturing a nearby causal SNP due to LD, we defined a true discovery as a SNP that either tags or is the causal SNP. Our simulation study showed that, if the LD regions are independent of the status of a SNP being truly null, then the functional *q*-value controls the FDR and the functional *p*-value controls the type I error rate. Finally, we have assumed that subjects in the primary study are not also included in the informative studies, so that the sets of *p*-values are independent under the null hypothesis.

A limiting factor for discovering genetic variants in a GWAS is the cost of acquiring samples. In this work, we demonstrate the utility of exploiting pleiotropy in a significance analysis for both small and large studies as a cost-effective strategy to increase power. While our emphasis is on leveraging pleiotropy from GWAS summary statistics, there is a large body of existing datasets to further increase statistical power, such as functional annotations in various cell types or states, expression-level data, or minor allele frequency. As such, we anticipate that sfFDR will have broader applications in genome-wide studies as a general framework that integrates informative data and provides a cost-effective way to improve power.

## Availability of data and materials

sfFDR is publicly available in the R package sffdr and can be downloaded at https://github.com/ajbass/sffdr. The code to reproduce the results in this work can be found at https://github.com/ajbass/sffdr_manuscript and the GWAS summary statistics used in the EGPA analysis are publicly available to download at https://www.ebi.ac.uk/gwas. Access to the UK Biobank data can be requested at https://www.ukbiobank.ac.uk/enable-your-research/apply-for-access.

## Data Availability

sfFDR is publicly available in the R package {\tt sffdr} and can be downloaded at https://github.com/ajbass/sffdr. The code to reproduce the results in this work can be found at https://github.com/ajbass/sffdr_manuscript and the GWAS summary statistics used in the EGPA analysis are publicly available to download at https://www.ebi.ac.uk/gwas.
Access to the UK Biobank data can be requested at https://www.ukbiobank.ac.uk/enable-your-research/apply-for-access.

https://www.ukbiobank.ac.uk/enable-your-research/apply-for-access

https://www.ebi.ac.uk/gwas

## Acknowledgements

This research has been conducted using the UK Biobank Resource under Applied Number 98032. This work was supported by the Wellcome Trust (WT220788, WT219506) and the MRC (MC_UU_00002/4, MC_UU_00040/01).

## 4 Methods

### 4.1 Overview

We first review the theory behind the functional false discovery rate (FDR) framework [15] and then introduce the functional *p*-value. Consider a GWAS study with *p*-values *P*_*i*_ for *i* = 1, 2, …, *m* SNPs. We initially assume that the *p*-values are approximately independent (via pruning or clumping) and identically distributed random variables (linkage disequilibrium considered in Section 4.3). The *p*-values follow a two group mixture model composed of SNPs that are not associated (i.e., null) with probability *π*_0_ or are associated (i.e., non-null or alternative) with probability 1 *− π*_0_. Let the status of SNP that is null be denoted by *H*_*i*_ = 0 and one that is non-null be denoted by *H*_*i*_ = 1. Suppose that there are *d* sets of informative GWAS summary statistics, ***Z***_*i*_ = (*Z*_*i*1_, *Z*_*i*2_, …, *Z*_*id*_), that can influence (i) the prior probability of a SNP being null, i.e., (*H* | ***Z*** = ***z***) *∼* Bernoulli(1 *− π*_0_(***z***)) and/or (ii) the distribution of the *p*-values under the alternative hypothesis, i.e., (*P* | *H* = 1, ***Z*** = ***z***) *∼ F*_1_(*·* | ***z***) where *F*_1_ is some distribution stochastically smaller than the Uniform distribution. Since we assume that individuals from the primary study are not in the informative studies, the summary statistics do not impact the *p*-values under the null hypothesis, i.e., (*P* | *H* = 0, ***Z*** = ***z***) = (*P* | *H* = 0) *∼* Uniform(0, 1). It is worth noting that the informative studies can share individuals between themselves.

Given the above assumptions, we can define a decision rule that incorporates the *p*-values and summary statistics to identify statistically significant SNPs. In particular, without loss of generality, we assume that the informative statistics are transformed to be uniformly distributed on the unit interval by using ranks. The significance region, Γ ∈ [0, 1]^1+*d*^, for the statistic *T* = (*P*, ***Z***) is defined as

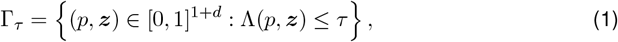

where *τ* ∈ [0, 1] is a significance threshold and

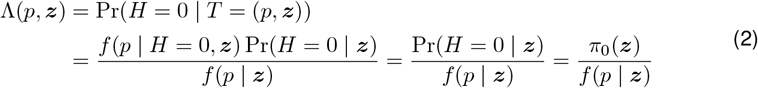

is the probability that a SNP is a false discovery given the observed data (i.e., the posterior error probability). Intuitively, the significance region classifies a set of SNPs with posterior error probabilities less than or equal to some threshold *τ* as “statistically significant.” The posterior error probability in this context is referred to as the functional local FDR and it is the optimal statistic for the Bayes rule with Bayes error [15]. As such, our strategy to optimally incorporate the summary statistic data is based on the functional local FDR.

Using the significance region in equation 1, we can construct the functional *q*-value (*q*_*f*_ - value) and *p*-value (*p*_*f*_ -value) which are different measures of significance for a SNP. Formally, the *q*_*f*_ -value is the minimum positive FDR (pFDR; closely related quantity to FDR) incurred when calling a SNP statistically significant [15, 18] while the *p*_*f*_ -value is the minimum type I error rate incurred when calling a SNP statistically significant. We note that these quantities have a Bayesian interpretation: the pFDR is the probability of a SNP being null given that it is classified as statistically significant [20], pFDR(Γ_*τ*_) = Pr(*H* = 0 | *T* ∈ Γ_*τ*_), and the type I error rate is the probability of a SNP being classified as statistically significant given that it is null, Pr(*T* ∈ Γ_*τ*_ | *H* = 0). Thus, for an observed statistic *t* = (*p*, ***z***), we can express the *q*_*f*_ -value as

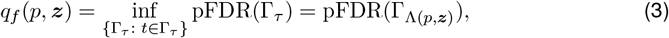

and the *p*_*f*_ -value as

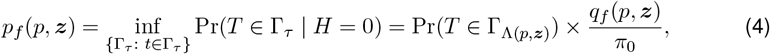

where 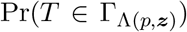 is the cumulative distribution function. While the definition of the *p*_*f*_ - value is the same as a standard *p*-value, we call it “functional” to emphasize that it is a function of the informative data.

The *q*_*f*_ -value and *p*_*f*_ -value are complementary quantities in a significance analysis: the former allows a researcher to decide the expected number of false discoveries they are willing to incur in the study while the latter allows for a standard *p*-value interpretation. We can use such measures of significance to identify statistically significant SNPs by either rejecting SNPs with a *p*_*f*_ -value below a significance threshold (e.g., 5 *×* 10^*−*8^) or a *q*_*f*_ -value below a desired FDR level. The mapping between the *q*_*f*_ -value and *p*_*f*_ -value provides different interpretations for the set of statistically significant SNPs, and thus connects a standard GWAS analysis to a FDR analysis while incorporating the informative data. In the next section, we discuss how to construct estimates of the functional local FDR, *q*_*f*_ -value, and *p*_*f*_ -value.

### 4.2 Estimating the functional local FDR, *q*_*f*_ -value, and *p*_*f*_ -value in the surrogate functional FDR framework

We first review construction of the *q*_*f*_ -value and *p*_*f*_ -value and then estimation in the surrogate functional FDR (sfFDR) framework. Given the significance region defined by equation 1, the *q*_*f*_ -value for the *i*th SNP is

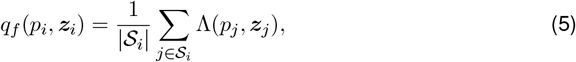

where 𝒮_*i*_ = {*j* : Λ(*p*_*j*_, ***z***_*j*_) *≤* Λ(*p*_*i*_, ***z***_*i*_)} is the set of SNPs with functional local FDRs below the value of the *i*th SNP [15]. The corresponding *p*_*f*_ -value is then

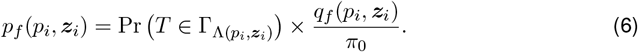

Since the *q*_*f*_ -value and *p*_*f*_ -value can be constructed from the functional local FDR 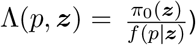, the primary quantities to estimate are *π*_0_ (***z***) and *f* (*p* ***z***).

The sfFDR framework provides estimates of the above quantities by extending the functional FDR framework to incorporate multiple GWAS summary statistics. In particular, we estimate *π*_0_(***z***) by minimizing the mean integral squared error using a generalized additive model (GAM) and *f* (*p* | ***z***) nonparametrically using a local likelihood kernel density estimator (KDE). We describe further details below and extend our discussion to include linkage disequilibrium in Section 4.3.

**Estimation of** *π*_0_(***z***) We extend the generalized additive model (GAM) method from ref. [15] to multiple informative variables. Let *η*_*λ*_(***z***) = **1**{*P>λ*|***Z***=***z***} denote a binary response variable where it follows that E[*η*_*λ*_(***z***)] = Pr(*P > λ* | ***Z*** = ***z***) *≥* Pr(*P > λ* | *H* = 0, ***Z*** = ***z***) Pr(*H* = 0 | ***Z*** = ***z***) = (1 *− λ*)*π*_0_(***z***) for some *λ* ∈ [0, 1). Given a set of informative variables, the general model is

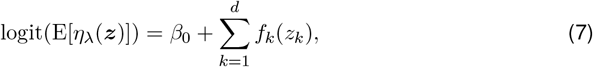

where 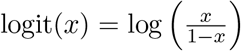, *β*_0_ is a constant, and *f*_*k*_(*z*_*k*_) is some function of the *k*th informa-tive variable. In this work, we use a natural cubic spline with knots chosen at specified quantiles (described below). Note that the above model allows for non-linear relationships and conservatively estimates the prior probabilities (or functional proportion of truly null hypotheses) at a given 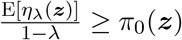.

We implement the following algorithm to estimate the functional proportion of truly null hypotheses. We first place the knots at regions that are likely to contain alternative *p*-values, i.e., the knots should be dispersed around small values (or lower quantiles) of *z*_*k*_. These regions will vary based on the signal density and power of the informative studies. We then fit the above model at *λ* = 0.05, 0.1, …, 0.9 and choose the fit that minimizes the mean squared integral squared error (MISE; see ref. [15]). The estimated functional proportion of truly null hypotheses at this minimum *λ* is 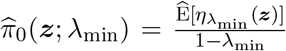, and, as discussed above, is con-servative. We note that if the test statistics are used (instead of *p*-values) then the knots should be placed where the signal is expected (i.e., the lower and/or upper tails of the distribution).

**Estimation of** *f* (*p* | ***z***) A challenge with nonparametric density estimation is that the joint density is difficult to estimate as the number of variables increases. We circumvent this difficulty by constructing a surrogate (or compressed) variable to reduce the dimensionality. In particular, we construct a surrogate variable based on 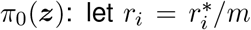 be the uniform quantile transformation of *π*_0_(***z***_*i*_) for *i* = 1, 2, …, *m*, where 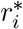 is the rank of the *i*th hypothesis (any ties are randomly assigned). We then estimate the density of 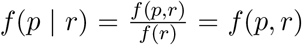 instead of *f* (*p* | ***z***), which is more tractable when there are many informative variables. To estimate *f* (*p, r*), we use a local likelihood KDE on the probit-transformed scale [15, 39]. The nearest neighbor smoothing parameter is chosen to be the estimated proportion of truly alternative tests of the *p*-values, i.e., the smoothing neighborhood covers 100 *×* (1 *− π*_0_)% of the data. Note that if 1 *− π*_0_ *<* 0.02 then we set the smoothing parameter to be 0.02.

In summary, we approximate the functional local FDR as

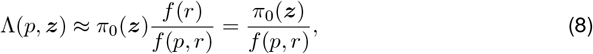

where the surrogate variable *r* is uniform quantile transformation of *π*_0_(***z***) and *f* (*r*) = 1. We refer to the above approximation as *surrogate* functional FDR (sfFDR) to emphasize that it is based on the surrogate variable *r*. Importantly, sfFDR reduces the dimensionality for tractable nonparametric density estimation. With the estimated (approximate) functional local FDR, we can then estimate the *q*_f_ -value and *p* _*f*_ -value as 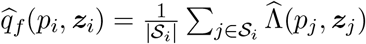 and 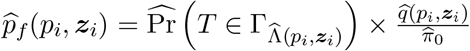, respectively. We note that 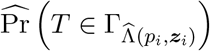 is the empirical CDF of the functional local FDRs and that the prior probability can be estimated as 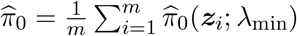 or using the maximum *q*_f_ -value in the study.

### 4.3 Extending the sfFDR framework to include SNPs in linkage disequilibrium

Thus far, we have assumed that a subset of SNPs have been selected to be approximately independent via pruning or clumping (i.e., no LD present). While this may be useful to understand a regions contribution to phenotypic variation, it is difficult to select the “best” representative SNP in an LD region. Therefore, we extend the sfFDR framework to circumvent such difficulty by providing a measure of significance for each SNP (including SNPs in LD) while incorporating the informative data.

To extend the sfFDR framework, we first model the proportion of truly null hypotheses, *π*_0_(***z***), and the joint density, *f* (*p, r*), on a set of LD-independent SNPs and then use the fitted curves to predict the corresponding values of the left-out SNPs (i.e., SNPs in LD; Figure 1). More specifically, we identify a subset of LD-independent SNPs via pruning, clumping, or by using the informative traits (see Section 4.7). Using the LD-independent SNPs, we apply the GAM method to estimate *π*_0_(***z***) and use the fitted curve to predict *π*_0_(***z***) of the left-out SNPs. After constructing the surrogate variable from the estimated *π*_0_(***z***), the joint density, *f* (*p, r*), and the marginal density, *f* (*r*), are estimated using the LD-independent SNPs. We note that the marginal density of the surrogate variable may not follow a uniform distribution when including SNPs in LD (i.e., *f* (*r*) ≠ 1). As such, we estimate the marginal density using a nonparametric KDE. Finally, the density values for the left-out SNPs are predicted from these fitted density curves. We can then estimate the functional local FDR 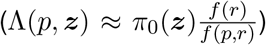 along with the corresponding *q*_*f*_ -value and *p*_*f*_ -value as outlined in Section 4.2.

### 4.4 Fine mapping with the functional local FDR

The FDR quantities estimated from the sfFDR framework can be used to perform fine mapping under the assumption that there is a single causal variant in a region [40]. More specifically, suppose there are *j* = 1, 2, …, *L* variants in a region of interest. The functional local Bayes’ factor (BF) can be expressed in terms of the functional local FDR as

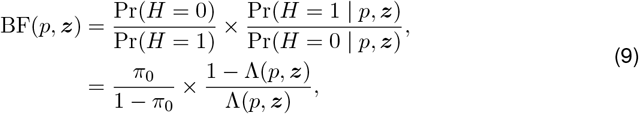

where *π*_0_ is the prior probability of the null hypothesis and Λ(*p*, ***z***) is the functional local FDR. Under the assumption of a single causal variant in the region, the posterior probability (PP) for the *i*th SNP is

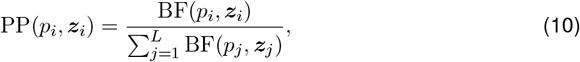

where we have implicitly assumed a uniform prior on any variant being the causal variant [40]. Therefore, the sfFDR framework provides estimates of the functional local BF and the corresponding PP for each SNP to help identify the causal locus. More generally, since the sfFDR framework incorporates SNP-level data, it is a novel framework to perform functional fine mapping. Note that the functional local BFs can also be used in any post-GWAS analysis in place of approximate BFs [31]. For example, while we do not explore it in this work, the functional local BFs estimated by sfFDR can also be used to perform colocalization [32, 33] while integrating informative data.

### 4.5 Simulation study

We conducted comprehensive simulations to assess the performance of the sfFDR framework. We simulated 150,000 independent hypotheses for the primary study with corresponding summary statistics for *k* = 1, 2, 3 informative studies. The proportion of null hypotheses was simulated as 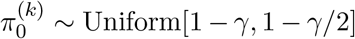, where the first 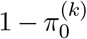 *p*-values were generated from the alternative distribution and the remaining were generated from the null distribution (standard Uniform distribution). We fixed the number of shared tests from the alternative hypothesis (i.e., overlap) between the informative studies and our primary study to be *γ* = 0.025 (a low level of overlap). Under the alternative hypothesis, we assumed the *p*-values followed a Beta(*α*, 5), where *α* = 2 for the “High” signal strength (or density), *α* = 3 for the “Medium” signal strength, and *α* = 4 for the “Low” signal strength cases. We describe below how the informative studies *p*-values (denoted by ***z*** = (*z*_1_, *z*_2_, *z*_3_)) influenced the prior probability *π*_0_(***z***) and the alternative density *f*_1_(*p* | ***z***) of the primary study *p*-values.

**Prior probability** *π*_0_(***z***) The relationship between the probability of a hypothesis test being truly null and the informative summary statistics was generated as follows. Define the function

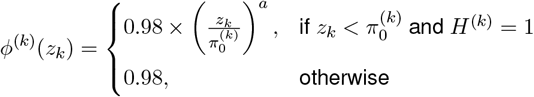

where *a* = 0.6 for the “Large” effect size strength case, *a* = 0.3 for the “Moderate” effect size strength case, and *H*^(*k*)^ = 1 for a test that is truly alternative in the *k*th informative study. The average of these components are then used to construct the prior probability of a hypothesis being null,

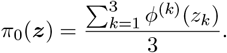

This relationship reflects the expected behavior where the prior probability decreases as the informative *p*-value decreases for shared alternatives. Using the prior probabilities, we then draw the true status of the *i* = 1, 2, …, *m* hypotheses as (*H*_*i*_ | ***Z***_*i*_ = ***z***_*i*_) *∼* Bernoulli(1 *− π*_0_(***z***_*i*_)). Under the null hypothesis (i.e., *H* = 0), the *p*-values follow a standard uniform distribution, i.e., (*P* | *H* = 0, ***Z*** = ***z***) *∼* Uniform(0, 1). We describe the distribution under the alternative hypothesis (i.e., *H* = 1) below.

**Alternative density** *f*_1_(*p* | ***z***) Under the alternative hypothesis, the distribution of the *p*-values depends on the informative variables. In particular, define the function

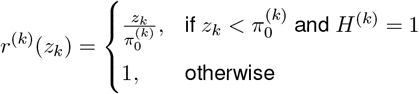

and

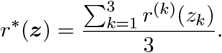

We assumed that the *p*-values follow a beta distribution under the alternative hypothesis, i.e., (*P* | *H* = 1, ***Z*** = ***z***) *∼* Beta(*α*(***z***), 5), where *α*(***z***) = *α*_0_ *− c ×* (1 *− r*^*∗*^(***z***)). The parameter *α*_0_ controls the signal strength (or density) of the alternative distribution and the parameter *c* controls the effect size strength of the informative summary statistics. We considered *α*_0_ = 0.3 for the “High” signal strength, *α*_0_ = 0.4 for the “Medium” signal strength, and *α*_0_ = 0.5 for the “Low” signal strength cases. The parameter *c* = *α*_0_*/*2 when the informative studies have a “Large” effect size strength and *c* = *α*_0_*/*4 when the informative studies have a “Moderate” effect size strength.

In total, there were 500 replicates at each combination of primary study signal strength, informative study signal strength, and the effect size strength of the informative studies. We also considered the scenario where the informative summary statistics have no impact on the primary *p*-values and so *π*_0_(***z***) = 0.98 and *α* = *α*_0_. For the *p*_*f*_ -values, we evaluated the type I error rate at a threshold of 1 *×* 10^*−*4^ and the power at 5 *×* 10^*−*8^. For the *q*_*f*_ -values, we evaluated the FDR at level 0.01 and the accuracy of the estimated proportion of truly null tests. We then compared the sfFDR framework to three different FDR procedures that can incorporate informative variables, namely, AdaPT [14], CAMT [16], and an estimator by Boca et al. (2018; referred to as the “Boca-Leek” method) [13]. The default settings of each software were used where the inputs were standardized (i.e., the informative summary statistics and the design matrix for the prior probability) across implementations. To assess our method under LD, for the *i* = 1, 2, …, *m* independent tests, we replicated the *p*-value and corresponding informative summary statistics *s*_*i*_ times, where *s*_*i*_ is drawn from the empirical distribution of the LD block sizes estimated using the UK Biobank (see Section 4.6). This reflects an extreme scenario where the SNPs in LD are perfectly correlated.

### 4.6 UK Biobank study

The UK Biobank is a repository of genetic, lifestyle, and health information for over half a million UK participants [41, 42]. Our analysis used four obesity-related traits that were rankbased inverse normal transformed, namely, body mass index (BMI), body fat percentage (BFP), cholesterol, and triglycerides. We restricted our analysis to 380,600 unrelated individuals with British ancestry. We then split the UK Biobank into two equal parts of size 190,300 where one part was used as the “primary” study and the other was the “informative” study.

Our trait of interest in the primary study was BMI and the informative traits were BFP, cholesterol, and triglycerides. We downsampled the primary study to 10%, 20%, …, 90%, 100% of the original sample size to study the impact of lower statistical power in our procedure. We applied the following processing to all downsampled datasets. Using the genotyped data (autosomes only), SNPs were filtered in PLINK with a MAF *<* 0.001, Hardy-Weinberg equilibrium *p*-value threshold of *<* 1 *×* 10^*−*10^, and a genotype missingness rate *>* 0.05. We then applied PLINK [43] for association testing while adjusting for sex, age, and the top 20 principal components provided by the UK Biobank to account for ancestry. Finally, we considered a “null” setting where the informative traits were permuted to be uncorrelated with BMI. In total, there were 10 permuted null datasets analyzed.

The set of LD-independent SNPs were determined by using pre-defined haplotype blocks constructed using the LDAK method [44]. More specifically, within each haplotype block, we performed hierarchical clustering using a random subset of 5,000 individuals from the UK Biobank to identify clusters of uncorrelated SNPs. In total, there were 161,207 “independent” clusters at a pruned correlation threshold of 0.99. At each cluster, we randomly selected a single representative SNP, and so the set of representative SNPs were approximately independent. We note that the LD-independent SNPs can be chosen other ways such as LD pruning or by using the informative traits (see Section 4.7). Finally, in our implementation of the GAM model, we fit a natural cubic spline to the informative traits *p*-values with knots placed at the 0.005, 0.025, 0.01, 0.05, 0.1 quantiles.

### 4.7 Application to EGPA study

We applied the sfFDR framework to a GWAS of eosinophilic granulomatosis with polyangiitis (EGPA). To illustrate our method on this rare disease, we used the *p*-values from a publicly available GWAS with 676 cases and 6,809 controls (see ref. [12] for analysis details). We note that, since the EGPA study only provided discrete *p*-values (2 significant digits), we recalculated the *p*-values using the publicly available effect sizes and standard errors and found a strong concordance with the published *p*-values (Figure S15). In total, there were 9,246,221 typed or imputed autosomal variants with INFO scores greater than 0.8 included in the analysis. The informative GWAS summary statistics were from clinically relevant features of EGPA, namely, childhood-onset asthma (13,962 cases and 300,671 controls) [23], adult-onset asthma (26,582 cases and 300,671 controls) [23], and eosinophil count (172,275 individuals) [24]. See the referenced publications for additional information on quality control steps. After removing SNPs in the MHC region and non-overlapping SNPs between EGPA and the informative traits, there were a total of 8,195,277 SNPs used in our analysis.

In our analysis, we considered 161,207 “independent” regions of the genome that were identified by a hierarchical clustering algorithm described in Section 4.6. To potentially increase the coverage of alternatives, we selected LD-independent SNPs as follows. For each region, if any of the informative traits *p*-values were below 0.001, then we selected the SNP that had the smallest *p*-value among the informative traits. Otherwise, we randomly selected a SNP in the region. When modeling the proportion of truly null hypotheses, we fit a natural cubic spline to the informative traits *p*-values with knots placed at the 0.005, 0.025, 0.01, 0.05, 0.1 quantiles. Finally, we used the original and permuted obesity-related traits (BFP, cholesterol, and triglycerides) from the UK Biobank (see Section 4.6) to assess whether sfFDR recovers the original *p*-values when the informative traits are unrelated to EGPA.

## 5 Supplementary materials

### 5.1 Supplementary tables

**Table S1:** The size of the 95% credible set using sfFDR and the standard (or original) *p*-values in the EGPA study. Note that only *TSLP* and *BCL2L11* are below genome-wide significance level for the standard *p*-values.

| Gene | sfFDR | Standard |
| --- | --- | --- |
| *BACH2* | 66 | 167 |
| *BCL2L11* | 13 | 14 |
| *GATA3* | 112 | 272 |
| *IKZF4* | 45 | 1,113 |
| *IRF1* | 52 | 39 |
| *LRRC32* | 136 | 2,564 |
| *RUNX1* | 141 | 68 |
| *TPRG1* | 45 | 64 |
| *TSLP* | 1 | 1 |
| *ZNF652* | 100 | 1,093 |

**Table S2:** The proportion of SNPs in the 95% credible set from sfFDR that overlap with the credible sets from the informative studies (ASTAO, ASTCO, and EOSC).

| Gene | ASTAO | ASTCO | EOSC |
| --- | --- | --- | --- |
| *BACH2* | 0.196 | 0.205 | 0.153 |
| *BCL2L11* | 0.968 | 0.000 | 0.850 |
| *GATA3* | 0.254 | 0.058 | 0.226 |
| *IKZF4* | 0.194 | 0.343 | 0.791 |
| *IRF1* | 0.320 | 0.000 | 0.000 |
| *LRRC32* | 0.131 | 0.161 | 0.112 |
| *RUNX1* | 0.076 | 0.081 | 0.000 |
| *TPRG1* | 0.173 | 0.000 | 0.822 |
| *TSLP* | 0.000 | 0.963 | 0.963 |
| *ZNF652* | 0.567 | 0.258 | 0.439 |

### 5.2 Supplementary figures

**Figure S1:**
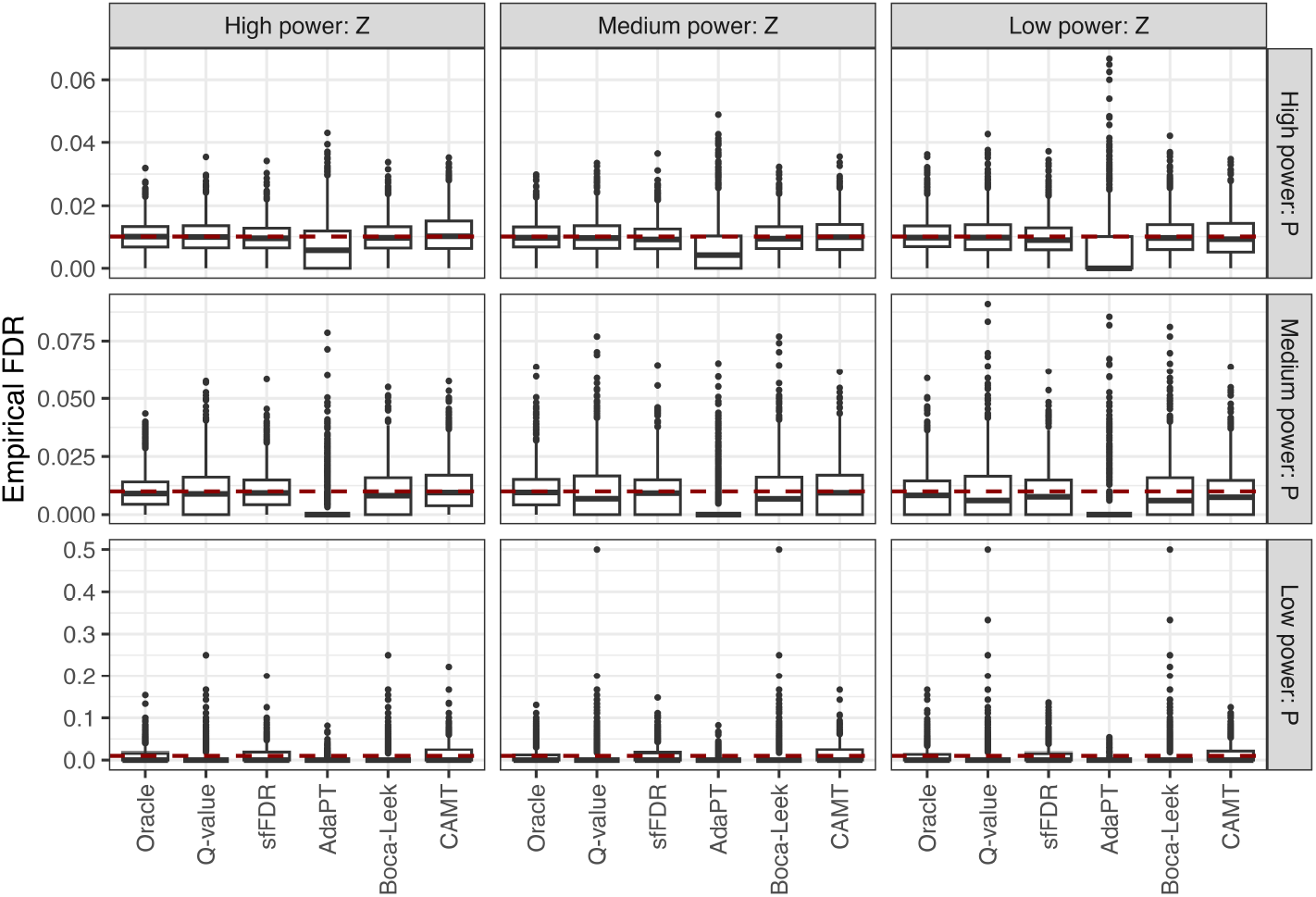
Assessing the target FDR at level 0.01 using the oracle functional *q*-values, standard *q*-values, functional *q*-values from sfFDR, Adapt, CAMT, and Boca-Leek in the independent SNP setting. The boxplot combines the “None,” “Moderate,” and “Large” effect size strength settings.

**Figure S2:**
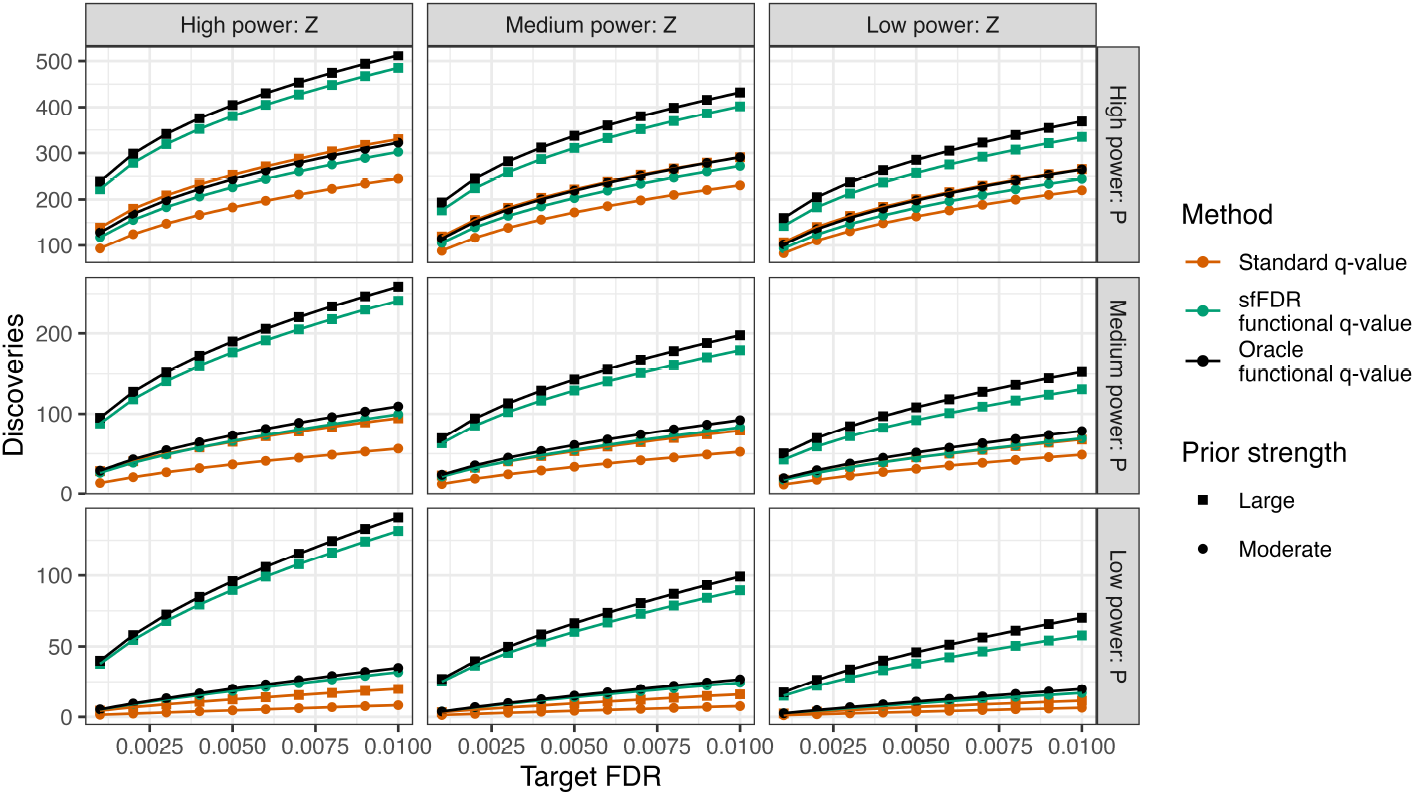
The number of discoveries as a function of the target false discovery rate (FDR) in the independent SNP simulation study using the standard *q*-value (dark orange), functional *q*-value from sfFDR (green), and the oracle functional *q*-value (black). We varied the power of the primary study (rows), the power of the informative studies (columns), and the effect size strength of the informative studies (shape). Each point is the average from 500 replicates.

**Figure S3:**
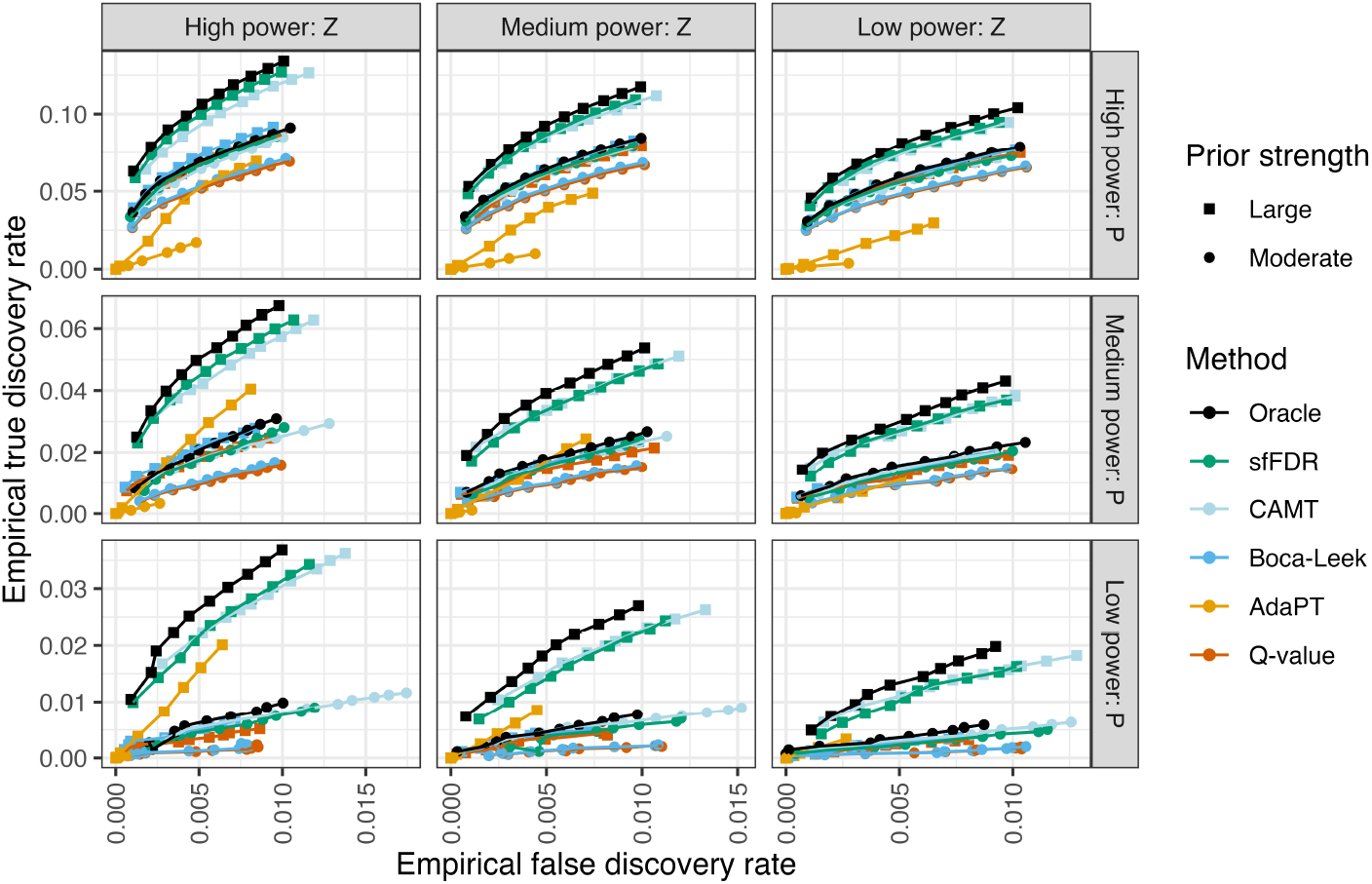
The empirical true discovery rate as a function of the empirical false discovery rate at a target FDR level of 0.001, 0.002, …, 0.01 in the independent SNP simulation study using the oracle functional *q*-value (black), functional *q*-value from sfFDR (green), CAMT (light blue), Boca-Leek (blue), AdaPT (orange), and standard *q*-value (dark orange). We varied the power of the primary study (rows), the power of the informative studies (columns), and the effect size strength of the informative studies (shape). Each point is the average from 500 replicates.

**Figure S4:**
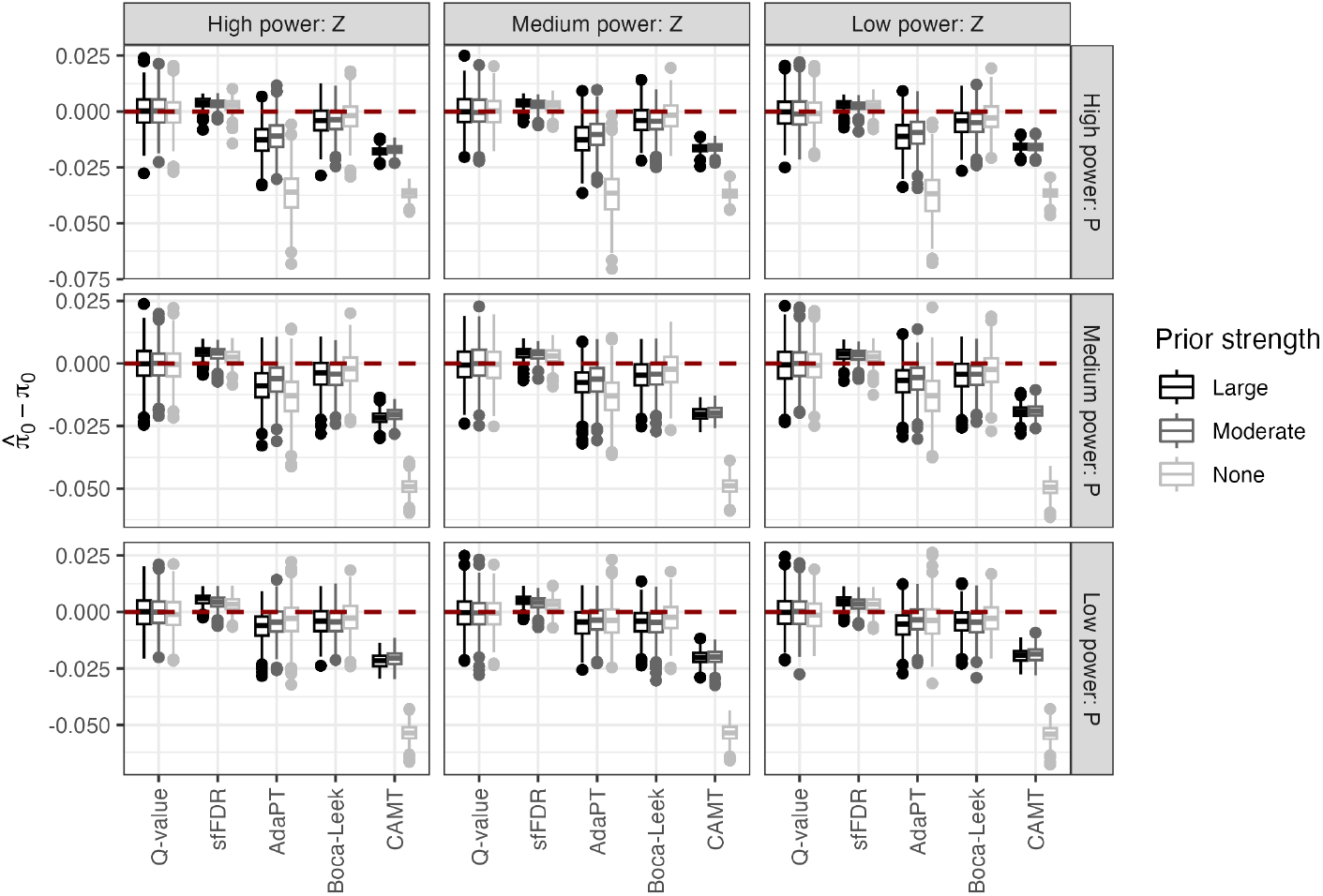
Comparing the estimated proportion of truly null tests from the standard *q*-value, sfFDR, Adapt, CAMT, and Boca-Leek in the independent SNP setting. There were a total of 500 replicates at each combination of primary study power (rows), informative study power (columns), and the effect size strength of the informative studies (color).

**Figure S5:**
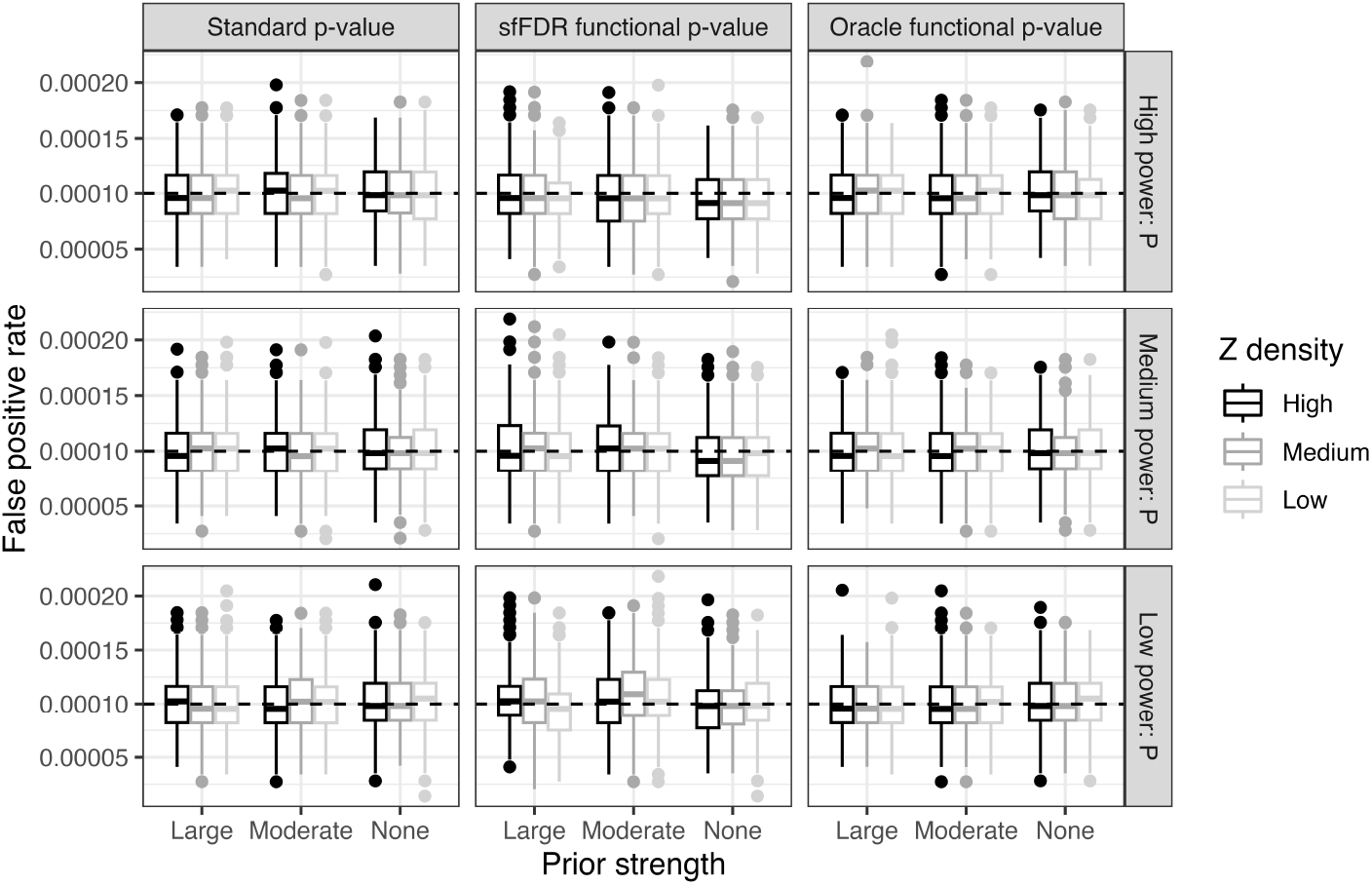
The type I error rate of the standard *p*-values, functional *p*-values from sfFDR, and the oracle functional *p*-values at a significance threshold of 1 *×* 10^*−*4^ in the independent SNP setting. We varied the power of the primary study (rows), the power of the informative studies (color), and the effect size strength of the informative studies (x-axis). There were a total of 500 simulations at each setting.

**Figure S6:**
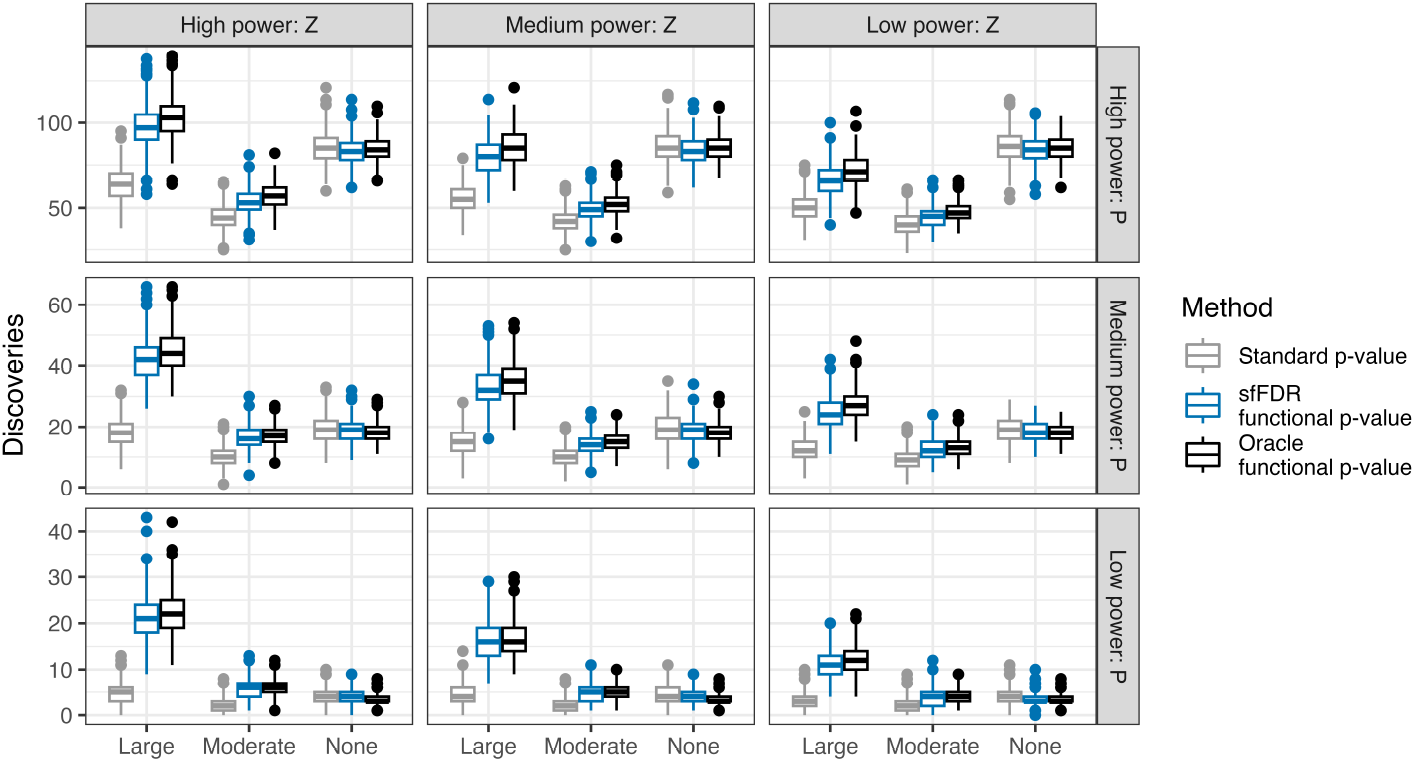
A comparison of the number of discoveries in the independent SNP simulation study using the standard *p*-value (grey), functional *p*-value from sfFDR (blue), and oracle functional *p*-value (black) at a significance threshold of 5 *×* 10^*−*8^. We varied the power of the primary study (rows), the power of the informative studies (columns), and the effect size strength of the informative studies (x-axis). There were a total of 500 simulations at each setting.

**Figure S7:**
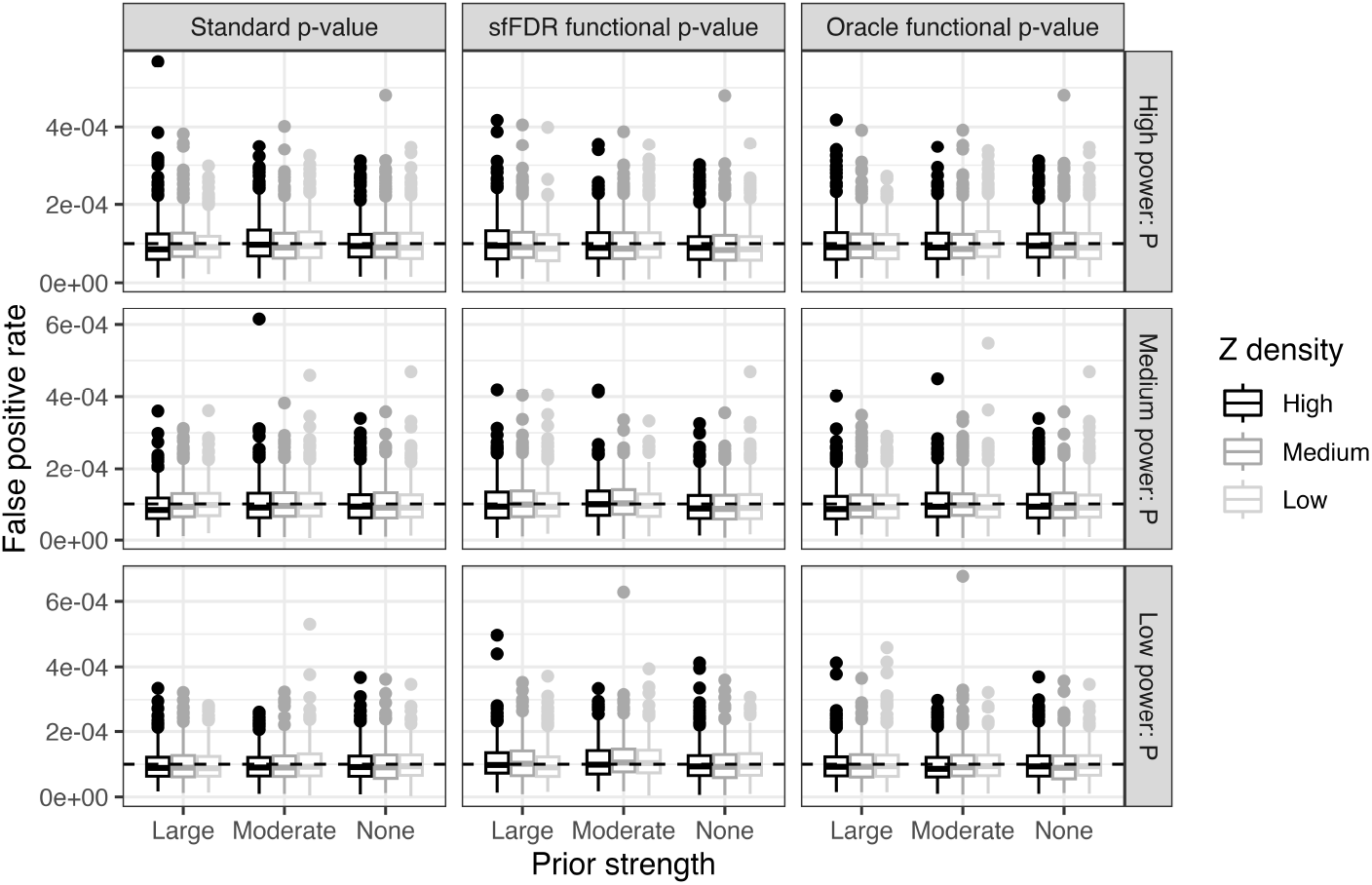
The type I error rate of *p*-values, functional *p*-values, and the oracle functional *p*-values at a significance threshold of 1 *×* 10^*−*4^ in the dependent SNP setting. We varied the power of the primary study (rows), the power of the informative studies (color), and the effect size strength of the informative studies (x-axis). There were a total of 500 simulations at each setting.

**Figure S8:**
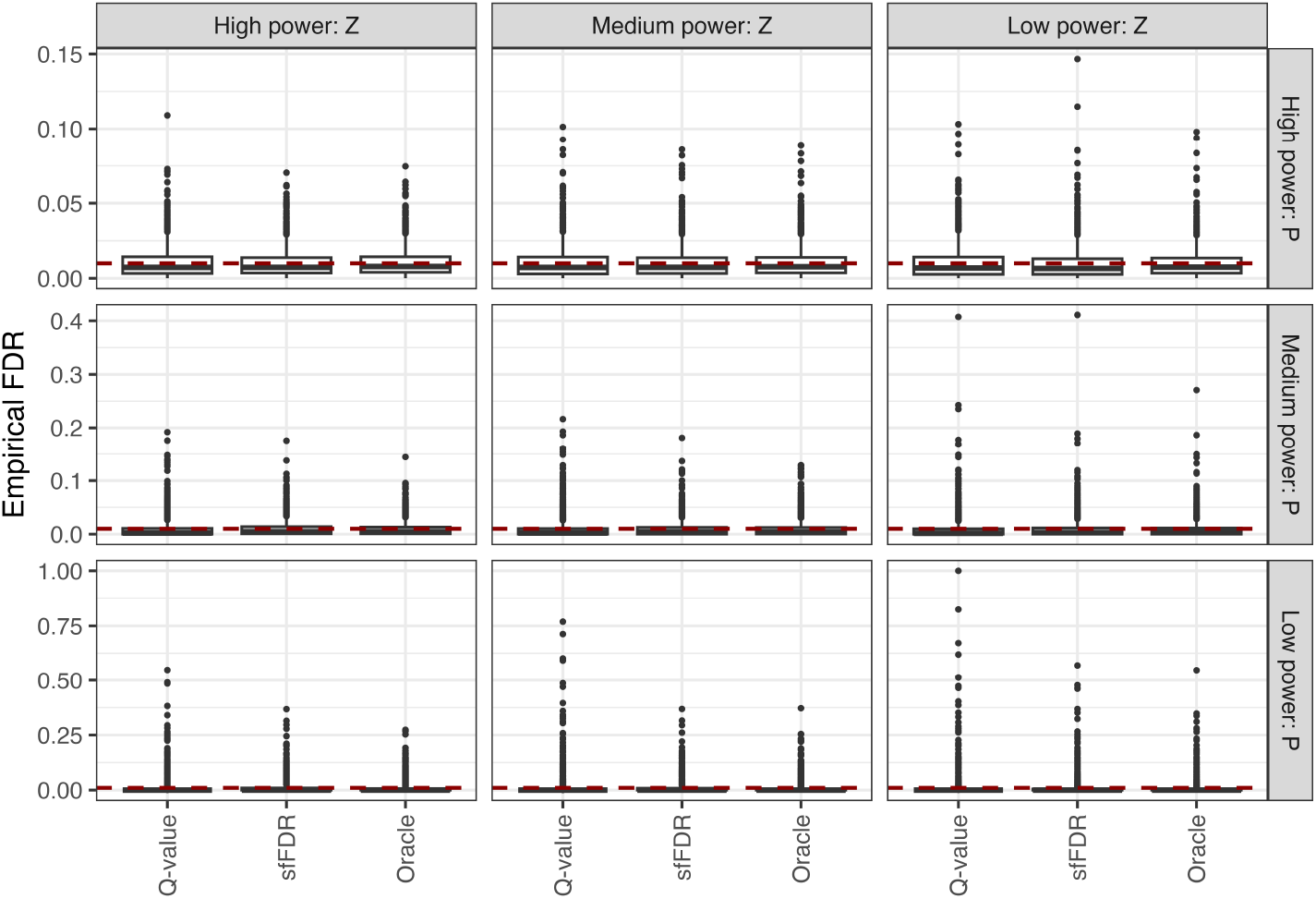
Assessing the target FDR at level 0.01 using the oracle functional *q*-value, standard *q*-values, and functional *q*-values from sfFDR in the dependent SNP simulation study. The boxplot combines the “None,” “Moderate,” and “Large” effect size strength settings.

**Figure S9:**
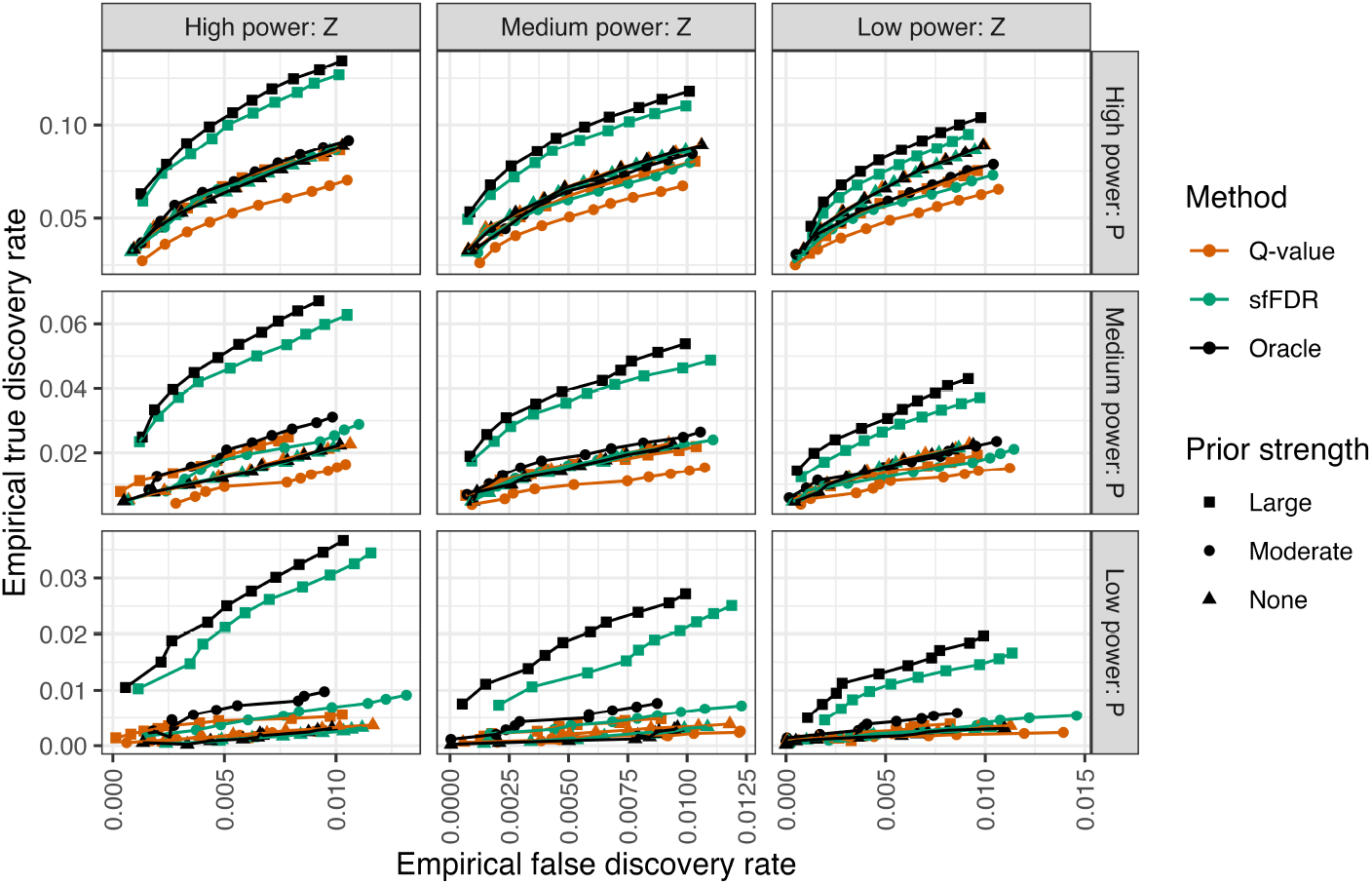
The empirical true discovery rate as a function of the empirical false discovery rate at target FDR level of 0.001, 0.002, …, 0.01 in the dependent SNP simulation study using the standard *q*-value (dark orange), functional *q*-value from sfFDR (green), and oracle functional *q*-value (black). We varied the power of the primary study (rows), the power of the informative studies (columns), and the effect size strength of the informative study (shape). Each point is the average from 500 replicates.

**Figure S10:**
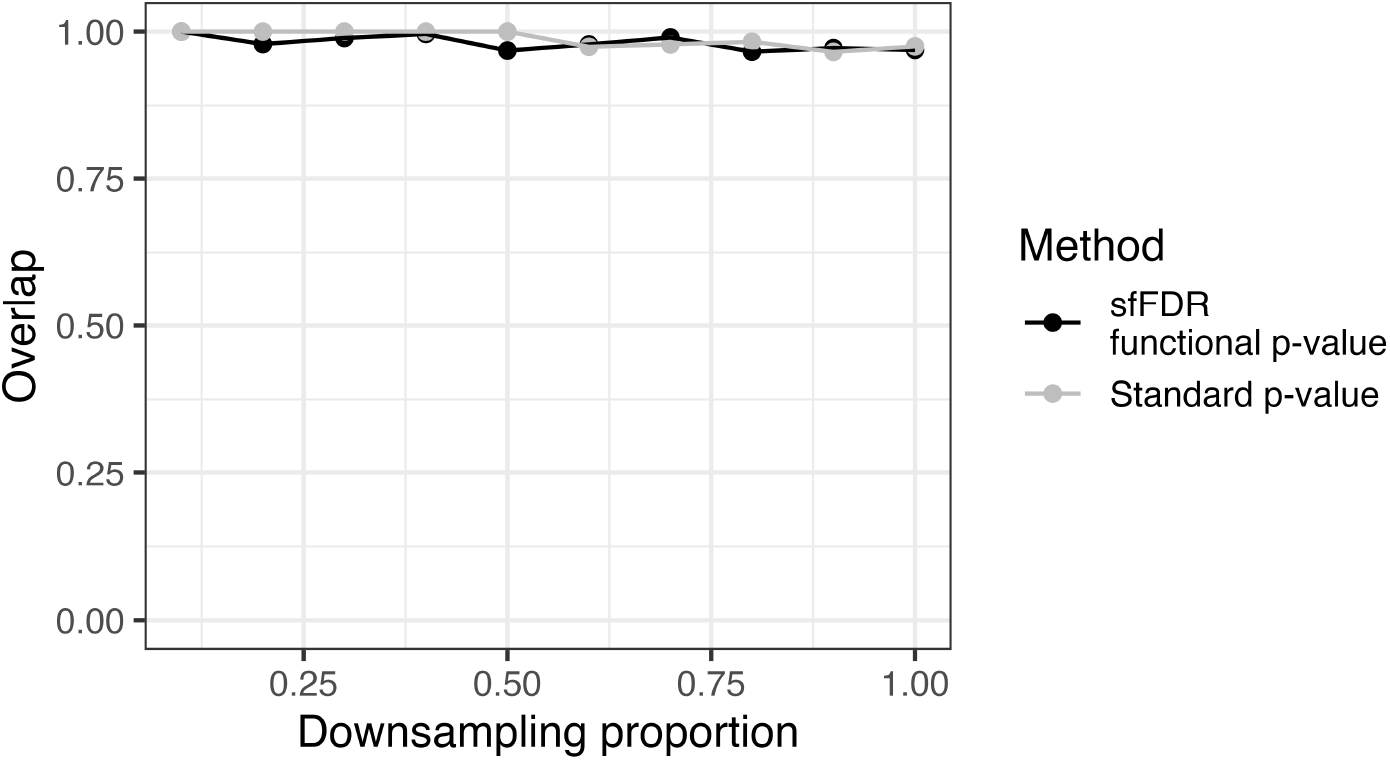
In the UK Biobank study, we performed a meta-analysis by combining the downsampled study (x-axis) with the informative study. At each downsampling proportion, we then calculated the proportion of discoveries from the functional *p*-value from sfFDR (black) and the standard *p*-values (grey) that overlapped with the meta-analysis at a significance threshold of 5 *×* 10^*−*8^.

**Figure S11:**
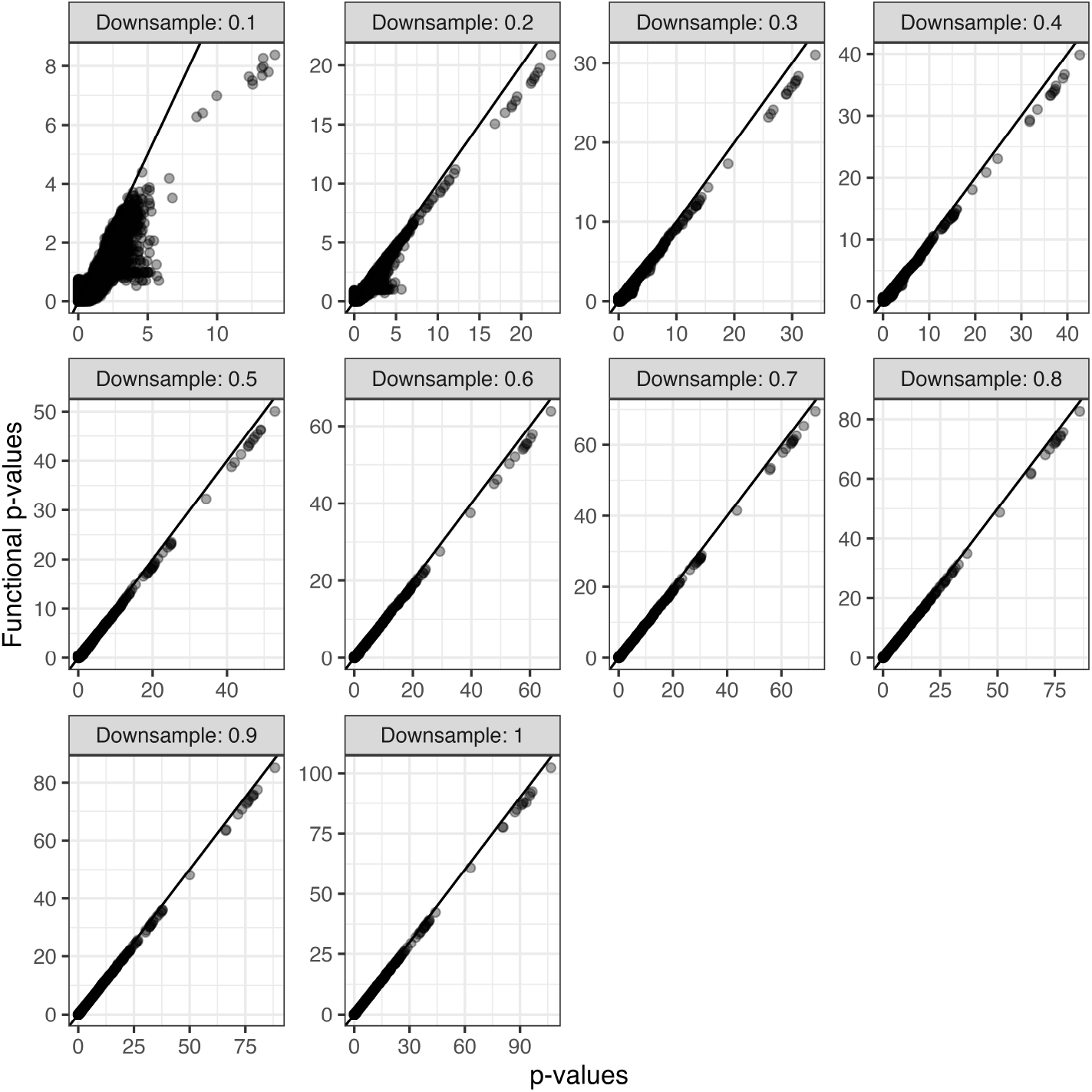
Comparison of functional *p*-values from sfFDR (y-axis) to UK Biobank standard *p*-values (x-axis) for BMI using a set of null correlated traits (body fat percentage, cholesterol, and triglycerides) as informative studies. There were 10 permutations of the null traits at each down-sampling proportion and each point represents the average functional *p*-value across permutations. A log10 transformation was applied to both axes.

**Figure S12:**
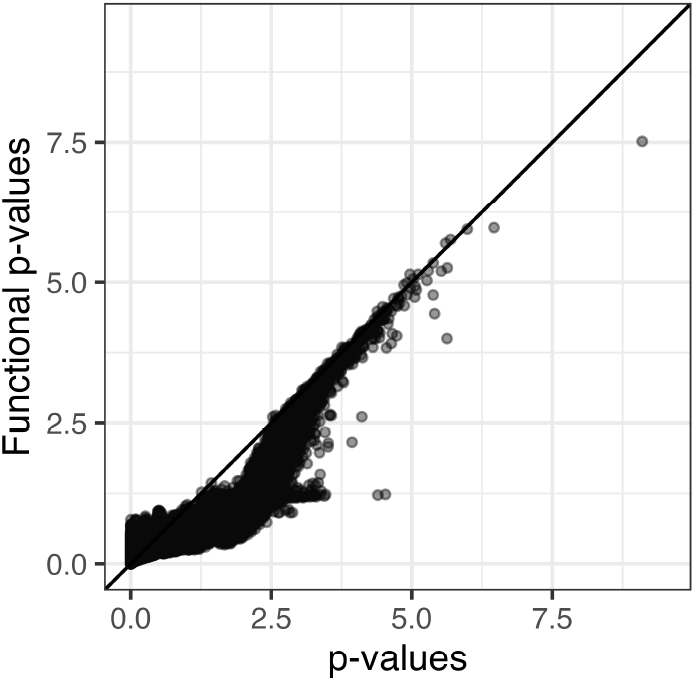
Comparison of functional *p*-values from sfFDR to standard *p*-values for the EGPA study when the informative traits are the UK Biobank null traits. Each point is the average of apply sfFDR to 10 replicates. A log10 transformation was applied to both axes.

**Figure S13:**
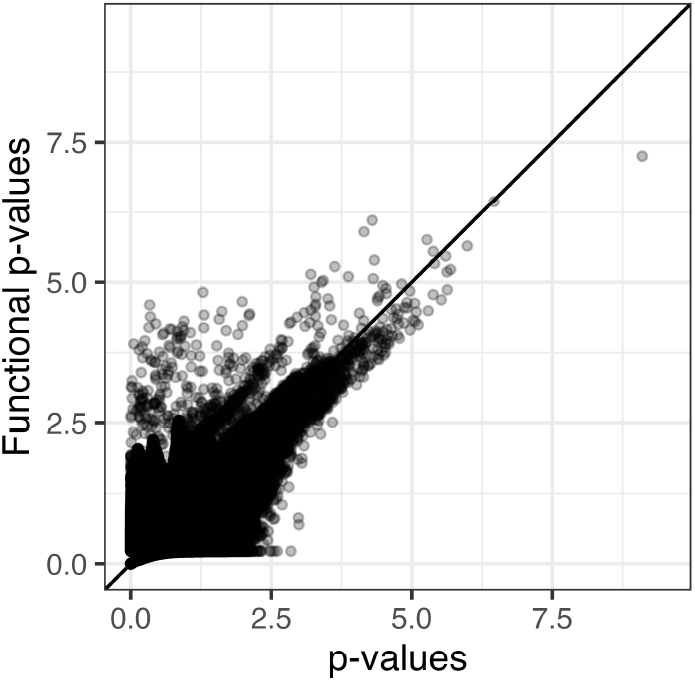
Comparison of functional *p*-values from sfFDR to standard *p*-values for the EGPA study when the informative traits are the UK Biobank obesity-related traits. A log10 transformation was applied to both axes.

**Figure S14:**
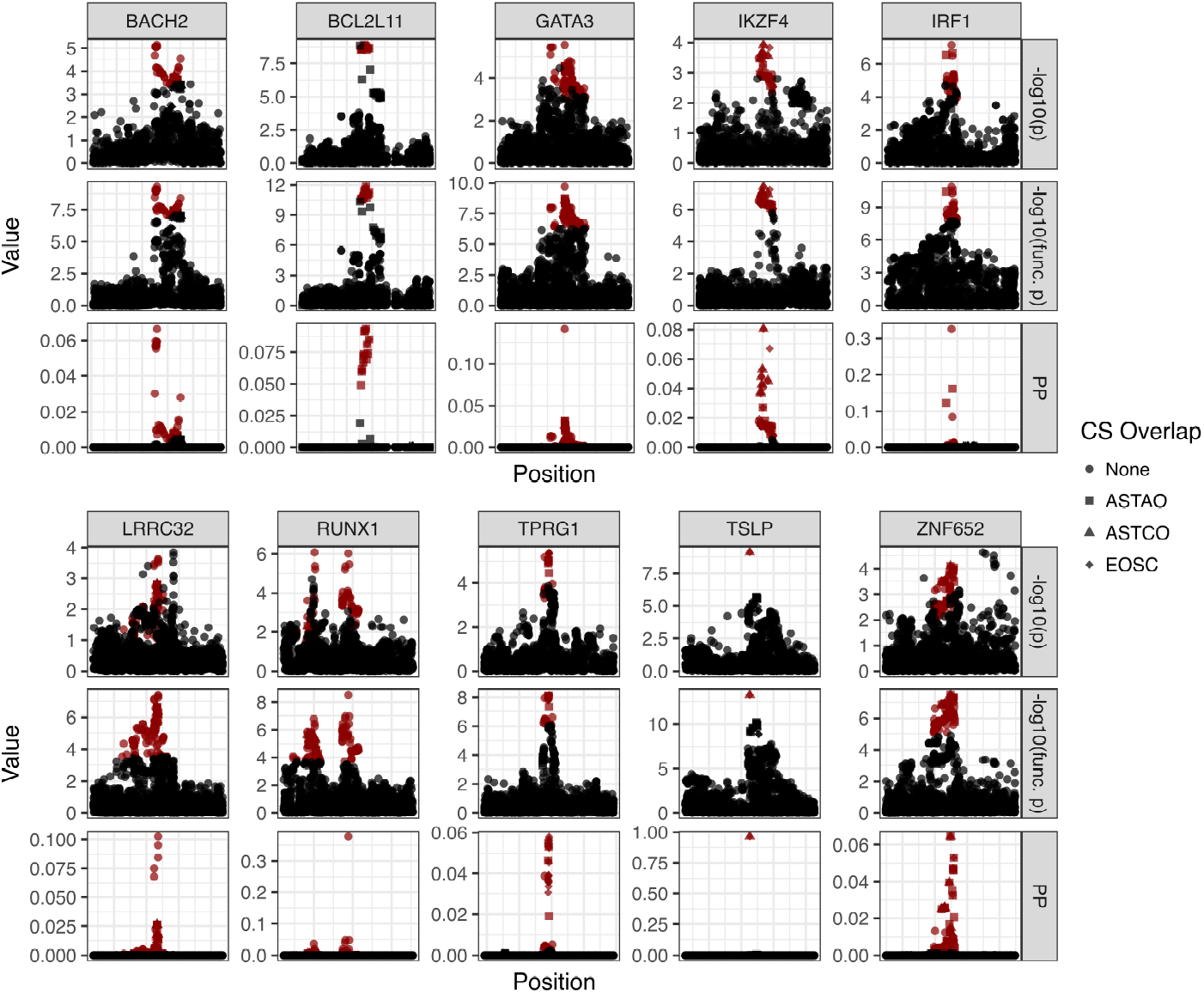
Fine mapping results using the functional local FDR estimates from sfFDR. For each lead SNP, the 95% credible set (CS) is shown in red for EGPA including SNPs 500kb upstream and downstream of the lead SNPs. The top plot in each set shows the local Manhattan plot while the bottom plot shows the fine mapping posterior probabilities calculated under the assumption of a single causal variant. We distinguish the SNPs in the CS that also overlap with the CS from ASTAO (square), ASTCO (triangle), and EOSC (diamond).

**Figure S15:**
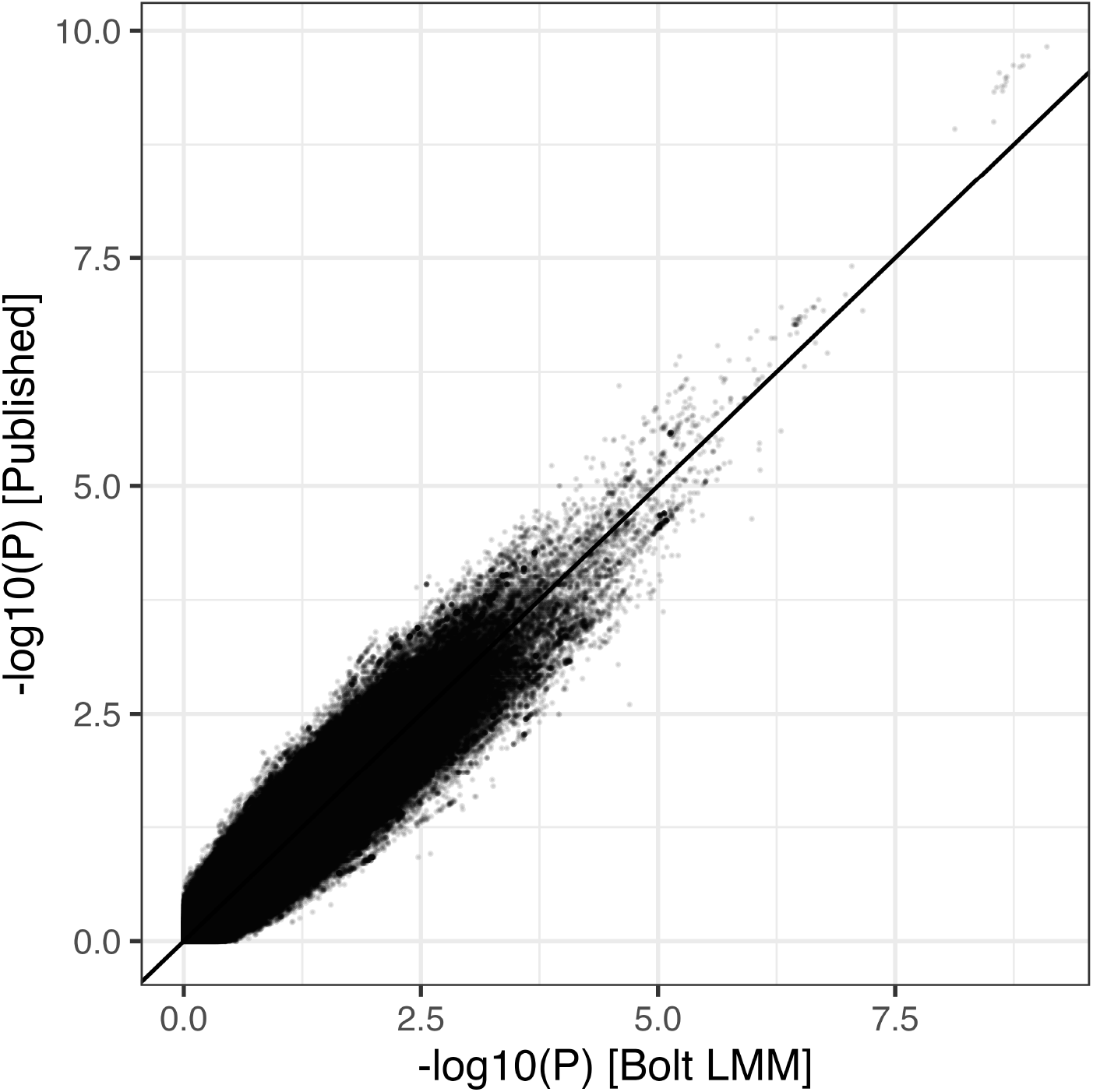
Comparison of the published *p*-values from the EGPA study and the *p*-values used in sfFDR.

